# Tau pathological binding activity in plasma before the onset of symptomatic Alzheimer’s disease

**DOI:** 10.64898/2026.04.03.26350110

**Authors:** Bernard J. Hanseeuw, Lisa Quenon, Jean-Louis Bayart, Emilien Boyer, Lise Colmant, Yasmine Salman, Thomas Gérard, Lara Huyghe, Vincent Malotaux, Pascal Kienlen-Campard, Flo Blondiaux Pirson, Renaud Lhommel, Laurence Dricot, Adrian Ivanoiu, Kavya Shamsundar, Winnie Pak, Joshua Soldo, Khalid Iqbal

## Abstract

Alzheimer’s disease (AD) and other tauopathies are characterized by the hyperphosphorylation of tau (pTau), leading to its aggregation in the brain, a process strongly predictive of neurodegeneration and cognitive decline. Currently, tau positron emission tomography (PET) is the only validated method for detecting tau aggregates in vivo. However, its high cost, invasiveness, and limited accessibility restrict its use in clinical settings and preclude large-scale screening. Moreover, existing plasma biomarkers that quantify the level of pTau at specific sites (e.g., pTau217) have limited specificity for confirming AD-related tau aggregation. There is therefore an urgent need for a reliable and scalable blood-based biomarker of tau pathology. A key mechanism underlying AD tau pathology is the ability of pTau to bind to and seed normal tau, facilitating prion-like propagation of insoluble tau aggregates. Here, we assessed the diagnostic performance of the VeraBIND Tau plasma assay, the first functional assay to detect pTau binding to recombinant normal tau using a tagged-tau chemiluminescent readout.

Seventy-nine cognitively unimpaired (CU) and 66 cognitively impaired older adults underwent blood sampling, cognitive assessment, amyloid-PET or cerebrospinal fluid analysis, and [^18^F]-MK6240 tau-PET imaging. Plasma pTau217 concentration was also quantified using the Lumipulse platform (Fujirebio).

VeraBIND Tau demonstrated 94.2% sensitivity and 96.1% specificity for predicting tau-PET positivity (AUC=0.97). It outperformed plasma pTau217 in CU individuals (PPV=85.9%), regardless of the pTau217 threshold used (maximal PPV of 57.5% using the 0.256pg/mL pTau217 threshold). This higher VeraBIND Tau diagnostic accuracy was driven by early tau-PET stages (Braak-like tau-PET stages 1-3; AUC=0.96 vs. 0.74 for pTau217, p=0.003). Both cross-sectional values and annual changes in VeraBIND Tau were associated with cognitive performance and entorhinal tau-PET signal (all absolute r_s_≥0.23, p<0.05). Combined with pTau217, VeraBIND Tau achieved 100% sensitivity and specificity for presymptomatic AD.

These findings highlight the strong promise of VeraBIND Tau as a scalable and accurate screening tool to detect AD tau pathology in the general population. Future studies should further evaluate its potential to optimize clinical trial enrichment, streamline diagnostic workflows, and support monitoring of tau-targeted therapies.

## Introduction

Alzheimer’s disease (AD) is biologically defined by amyloid-beta (Aβ) plaques and neurofibrillary tau tangles (NFTs), which can be observed *in vivo* using positron emission tomography (PET) imaging.^1^ These pathologies develop silently, before overt cognitive decline can be detected, offering a unique opportunity to screen for AD pathology and prevent the disease from occurring.^2^ Previous studies showed that clinical impairment is closely associated with NFTs.^3,4^ Clinically unimpaired (CU) individuals with both elevated Aβ and tau-PET signals (A+T+) face a high risk of cognitive impairment within 3-5 years (risk=53-57%) compared to those with only high Aβ burden (A+T-, risk=8-17%) and negative individuals (A-T-, risk=3-6%).^5,6^ Identifying A+T+ individuals in the general population is key for prevention, but challenging as their prevalence is estimated at ∼10% of CU individuals at age 75.^6^ As PET imaging is invasive and not widely available, developing blood-based biomarkers reflecting the active formation of tau aggregates is critical for envisioning population screening.

Current blood-based biomarkers reflecting AD pathology include plasma levels of Aβ peptides (Aβ42/Aβ40 ratio)^7–9^, hyperphosphorylated tau (pTau) species at different sites (e.g., pTau181, pTau205, pTau212, pTau217, pTau231)^10–17^, and the pTau217/Aβ42 ratio^18–20^. Compelling evidence suggests that these plasma biomarkers, including pTau species, more closely reflect Aβ plaque load than tau pathology, as evidenced by PET.^14,21,22^ Among them, plasma pTau217 presents the widest dynamic range and best correlates with amyloid-PET imaging and post-mortem measures.^23–26^ Recent meta-analyses demonstrated excellent accuracy to predict Aβ and tau-PET positivity in clinically impaired (CI) individuals.^27,28^ However, the accuracy of pTau217 for detecting tau-PET positivity is low in CU individuals.^27,28^ Specifically, using high thresholds, plasma pTau217 can identify A+T+ individuals with advanced tau pathology (e.g., Braak-like tau-PET stages≥4)^29–32^, while using lower thresholds will identify both A+T+ and A+T-CU individuals. These observations indicate that plasma pTau217 may serve a diagnostic purpose in symptomatic individuals, as recently approved by the Food and Drug Administration, but lacks accuracy as a stand-alone screening test in CU individuals to detect incipient tau pathology.

Current plasma pTau species assays may lack specificity for capturing early aggregated tau pathology, as tau phosphorylation patterns are both overlapping and heterogeneous across physiological and pathological conditions.^33^ Tau contains many sites at which it is hyperphosphorylated reversibly under various stress conditions such as hypothermia. In addition, existing assays primarily quantify absolute concentrations of individual pTau species, making them vulnerable to interindividual variability. Factors such as chronic kidney disease and ancestry-related biological differences can alter circulating pTau levels independently of brain tau pathology, increasing the likelihood of false-positive or false-negative interpretations.^34–37^ Therefore, increased concentrations of tau hyperphosphorylation, especially at any single phosphorylation site, does not necessarily indicate the presence of tau pathology. One key pathological mechanism in AD and other tauopathies involves tau seeding activity whereby specific misfolded pTau species binds to and templates normal tau, promoting prion-like aggregation and spreading of tau pathology.^38–40^ In this work, we evaluated a novel functional plasma assay, named VeraBIND Tau, which reflects the binding activity of pTau as compared against [^18^F]MK6240 tau-PET imaging as the *in vivo* gold standard to detect tau pathology in the brain. This functional assay tests the pathological activity of pTau from plasma by measuring its ability to bind recombinant normal tau *in vitro*. The VeraBIND™ technology is a sample transformation and biomarker purification technology that purifies and enriches low-abundance pTau peptides from plasma for their subsequent characterization. The enriched pTau peptides are then incubated with recombinantly-tagged normal tau, and binding of this recombinant normal tau by pathologically active pTau (PA pTau) is detected with an anti-recombinant tag alkaline phosphatase antibody conjugate. The resulting relative luminescence unit (RLU) is directly proportional to the amount of recombinant normal tau bound by pTau in the plasma sample. The RLU signal is converted into a ratio using a Standard run in each assay, providing a semi-quantitative result. In this study, we assessed the diagnostic performance of VeraBIND Tau to predict tau-PET positivity in a mixed sample of CU and CI participants, as well as in each of these groups and in low versus high Braak-like tau-PET stages. Its diagnostic performance was compared to other blood-based biomarkers, including the plasma concentration of pTau217 and pTau181, which are currently the main blood-based biomarkers approved for clinical use. Moreover, we assessed the value of a two-test approach for the detection of A+T+ individuals by combining VeraBIND Tau, to reflect tau-PET positivity, with pTau217, to predict Aβ positivity. Finally, we investigated the associations between the VeraBIND Tau semi-quantitative measure, cognition, regional tau-PET signal, the Centiloid value, and other blood-based biomarkers, both cross-sectionally and longitudinally.

## Materials and methods

### Participants

In total, 145 individuals aged over 45 years old were included in this study. This sample was composed of 76 patients recruited at the Memory Clinic of the Cliniques Universitaires Saint-Luc in Brussels (Belgium) and 69 volunteers selected from a local academic study registry. Volunteers’ selection was enriched for carriers of the Apolipoprotein E ε4 allele (*APOE* ε4) to match the frequency of patients’ *APOE* ε4 carriership (see Supplementary Material for the *APOE* genotyping method). Recruitment and baseline examinations were conducted between June 2019 and April 2025.

Exclusion criteria were any neurological conditions associated with long-term risk of significant cognitive impairment or dementia (e.g., multiple sclerosis, Huntington’s disease), focal brain lesions, psychiatric disorders (e.g., major depression, schizophrenia, bipolar disorder), active alcohol and drug abuses, and participation in a clinical trial of an investigational product.

This study was approved by the Ethical Committee of UCLouvain (#UCL-2016-121, Date: 13/05/2019; Eudra-CT number: 2018-003473-94) and was conducted in compliance to the Declaration of Helsinki principles. Written informed consent was obtained from all participants.

At study inclusion, participants underwent blood sampling, anatomical brain magnetic resonance imaging (MRI), lumbar puncture, or an amyloid-PET, tau-PET imaging, and a comprehensive cognitive assessment. Longitudinal cognitive assessments are subsequently proposed every two years.

### Blood-based biomarkers

#### Blood draw and plasma preparation

A standard venipuncture procedure was performed using a 21g needle, and blood was collected in ethylenediaminetetraacetic acid (EDTA) polypropylene K2 tubes (K2-EDTA tubes, 7.5 mL S-monovette 01.1605.008, Sarstedt®). The tube was placed on ice immediately after collection and plasma isolation was performed within two hours. Blood was centrifuged at 2000×g for 10 min at 4°C. Extracted plasma was aliquoted at a volume of 500 μL into cryotubes then frozen within two hours of blood collection at −80°C until further analysis^7^. For cross-sectional analyses, we used the VeraBIND Tau scores closest in time to tau-PET (average delay= 0.4 years, SD=0.8). The delay between VeraBIND Tau scores, amyloid-PET and cognitive data was on average of 0.8 (SD=0.8) and 0.5 years (SD=0.6), respectively.

#### VeraBIND^TM^ Tau assay

The VeraBIND™ Tau plasma assay was developed using the Veravas VeraBIND™ (Biomarker Isolation and eNrichment for Detection) sample transformation and biomarker purification technology, and a pool of proprietary monoclonal antibody-coated capture beads to specifically capture and purify pTau from plasma (Fig.1). A detailed description of the equipment, reagents and procedures used to perform the assay is provided in the Supplementary Material.

**Fig. 1.**
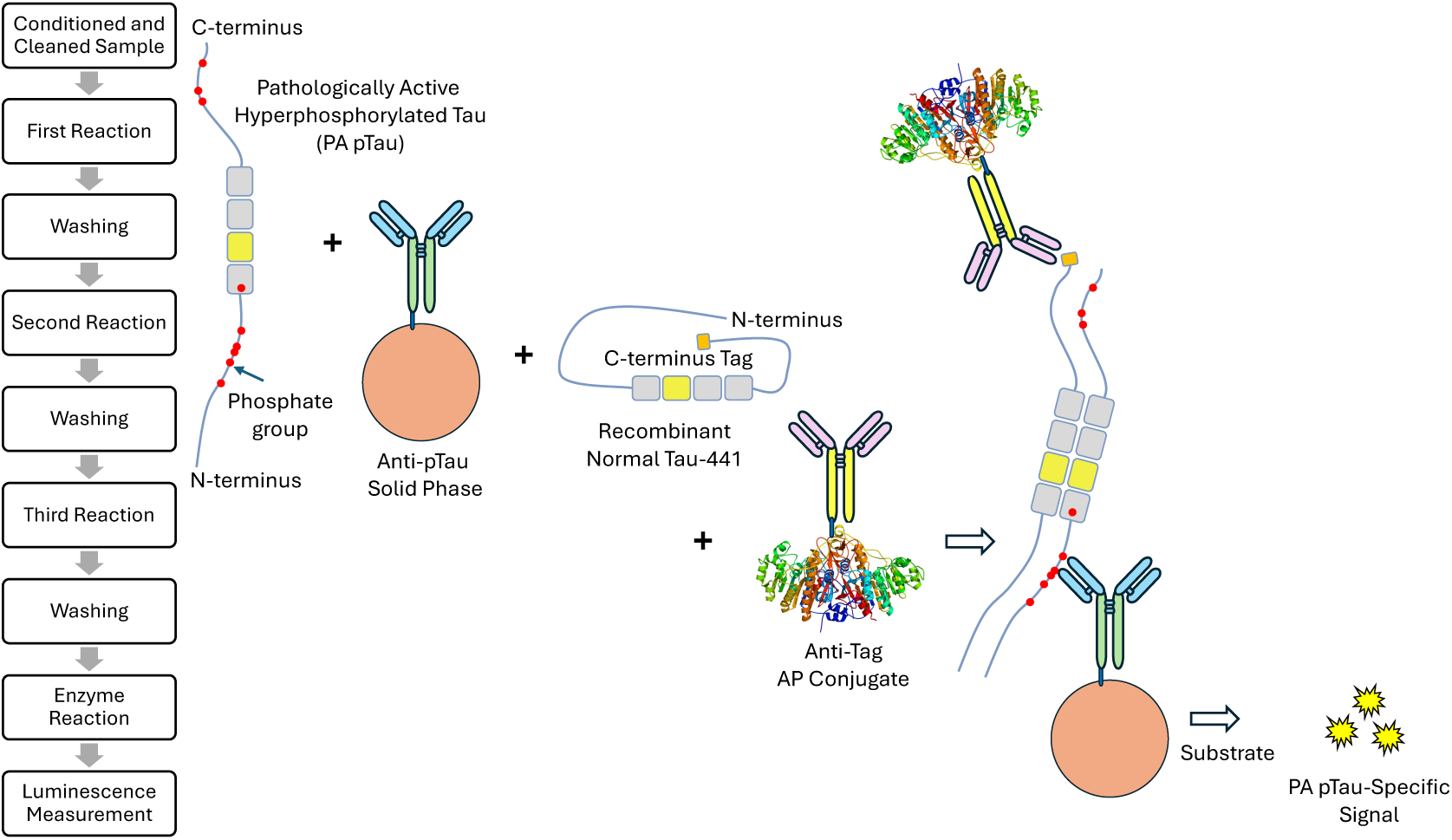
VeraBIND Tau assay. The VeraBIND Tau assay uses a pool of different antibody-coated capture beads to capture hyperphosphorylated tau (pTau) in the plasma sample. The capture beads are washed to remove the plasma matrix, and buffer-exchanged/washed into a normal Tau binding buffer that mimic the pH, ionic and hydrophobic binding environment in the brain. The assay adds recombinant full-length normal Tau 1-441 with a recombinant tag at the C-terminus (recombinant normal tau) to determine if any of the pTau captured on the beads is pathologically active (PA pTau). If pTau captured from the plasma is pathologically active, it will bind the recombinant normal tau like it does in the brain with tau pathology. The final step washes the capture beads to remove unbound normal tau, adds an alkaline phosphatase (AP) conjugated anti-tag monoclonal antibody (Anti-Tag AP Conjugate), washes the capture beads to remove unbound AP Conjugate, and adds Substrate to generate a relative luminescence signal (RLU) that is directly proportional to the amount of recombinant normal tau bound by pTau in the plasma sample.

After thawing the plasma sample at 2-8°C, diluting the sample to inactivate endogenous plasma phosphatases, and pre-analytical cleaning of the sample using VeraBIND clean beads, the pool of monoclonal antibody-coated capture beads are added for the selective capture and purification of pTau. The capture beads are magnetically washed, buffer exchanged, and mixed into normal tau binding buffer to facilitate ionic and hydrophobic binding of pTau to recombinant normal Tau (nTau). Next, recombinant V5-tagged full-length nTau 1-441 (Recombinant Normal Tau-441) is added, and incubated static overnight at 2-8°C for the binding of the Recombinant Normal Tau-441 by any pTau, akin to the binding of pTau to nTau that occur in the brain of AD patients. The next day, after the capture beads are washed to remove non-bound Recombinant Normal Tau-441, anti-V5 alkaline phosphatase (AP) Conjugate is added to detect any Recombinant Normal Tau-441 bound by pTau on the capture beads. After washing the capture beads to remove any non-bound AP Conjugate, the capture beads are mixed with substrate and incubated at 30°C for 60 min to generate a luminescence signal. The relative luminescence units (RLU) are directly proportional to the amount of Recombinant Normal Tau-441 bound by pTau. The semi-quantitative VeraBIND Tau assay results are reported as a test result Score which is calculated using an EDTA plasma-based Standard (e.g., healthy control plasma). A test result Score <1.0 is a negative test result, meaning that PA pTau has not been detected in the plasma sample, while a Score ≥1.0 is a positive test result, indicating that PA pTau has been detected in the plasma sample.

#### Quantification of plasma pTau217 levels

Following one hour room temperature thawing, pTau217 concentration was measured on a Lumipulse G600II analyzer, using the Lumipulse® G pTau217 Plasma RUO assay (Fujirebio, Ghent, Belgium).

#### Quantification of plasma pTau181, pTau231 and the Aβ42/40 ratio

Quantification of soluble pTau181, pTau231, Aβ40 and Aβ42 was performed using the SIMOA (Single-Molecular Array, Quanterix)^7,41^ pTau-181 Advantage V2.1 (REF: 104111), pTau-231 Advantage (REF: 102292), and Neurology Plex 3 A (REF:101995) kits. Each plasma sample was thawed to room temperature for one hour before being processed.

### Amyloid measurement

The amyloid status was determined using cerebrospinal fluid (CSF) Aβ42 concentrations (n=44; Lumipulse automated assay) or Αβ PET-scan (n=101) with [^18^F]Flutemetamol (Vizamyl^TM^, GE Healthcare) or [^11^C]Pittsburgh compound B (PiB). PET acquisition and reconstruction details are provided in the Supplementary Material.

For both radiotracers, semi-quantitative PET data were computed using PNEURO software (v4.1; PMOD LLC Technologies, Zurich, Switzerland) following the previously developed Centiloid pipeline^42^ to return a Centiloid value for each participant.

Participants were considered amyloid positive (A+) for a Centiloid value≥20^43^ or CSF Aβ42 levels≤544 pg/mL.^44,45^

### [^18^F]MK6240 tau-PET

#### Acquisition

[^18^F]MK6240 (Lantheus Inc.) is an investigational drug studied as a second-generation cerebral tau tangles imaging agent. Radiosynthesis was conducted at KULeuven (Leuven, Belgium) and delivered to our clinic in less than an hour. Ninety minutes after intravenous administration of [^18^F]MK6240 (target activity 185±5 MBq) a 30-min dynamic list-mode acquisition was performed on a Philips Vereos digital PET-CT (Philips Healthcare, Amsterdam, Netherlands). Images were reconstructed using the manufacturer’s reconstruction algorithm, which includes attenuation, scatter, and decay corrections, and time-of-flight information. Point spread function and 1 mm reslicing was also computed using the manufacturer’s algorithm to obtain a better resolution recovery.^46^

#### Visual Braak-like staging

Each tau-PET scan was co-registered with a 3D T1-weighted MRI (acquired using a Signa^TM^ Premier 3 Tesla head scanner - GE Healthcare, Chicago, USA - equipped with a 48-channel phased-array head coil; resolution: 1x1x1mm) to ensure precise anatomical localization. Two experienced nuclear physicians (R.L. and T.G.) determined a visual Braak-like stage (ranging from 0 to 6) by assigning to every participant the furthest Braak-like region-of-interest (ROI) where a significant signal was observed, excluding meningeal non-specific uptake. For some intricate scans, MRI segmentation was used to assist visual reading. Braak-like ROIs were defined based on previous work led by Schöll et al. on in vivo Braak staging using brain imaging^47^ (see Supplementary Material). The tau-PET status was considered positive (T+) for Braak-like stages>0.

#### Regional tau burden quantification

PET images were registered on T1-weighted structural MRI images using PetSurfer pipeline, a set of tools within FreeSurfer for end-to-end integrated MRI-PET analysis.^48,49^ Standardized Uptake Value ratio (SUVr) values were extracted for all regions from the Desikan-Killiany Atlas^50^, using the cerebellum gray matter as reference region. In this study, analyses focused on the bilateral entorhinal and inferior temporal cortex.

### Neuropsychological assessment

All participants underwent a neuropsychological testing that evaluated global cognition using the Mini-Mental State Examination (MMSE^51^) and four cognitive domains: verbal episodic memory (Free and Cued Selective Reminding Test, French version^52^), language (Lexis Naming Test, Category and Letter Fluency Test for animals and letter P^53^), executive functions (Trail Making Test part A and B^54^ and Luria’s Graphic Sequences^55^), and visuospatial functions (Clock Drawing Test^56^ and Praxis part of the CERAD battery^57^). *Z*-scores were computed for each cognitive domain based on three measures for each domain^58^, based on an independent sample composed of 32 clinically unimpaired (CU) individuals who remained cognitively stable over eight years. A cognitive domain was considered impaired if the z-score fell below -1.5. Participants were considered as being clinically impaired (CI) if at least one z-score was below this cut-off and as being CU otherwise.

### Statistical analysis

Statistical analyses were performed using R (2022.07.2). The alpha statistical significance threshold was set at 0.05. Statistical significance was uncorrected for multiple comparisons, otherwise stated.

#### Participants’ characteristics

Group differences in demographics, biomarkers and cognitive measures were assessed using independent-sample t-test when the assumptions of normality and homoscedasticity were satisfied, and Mann-Whitney U test otherwise. Chi-squared tests were used to examine differences in *APOE* ε4 carriership, sex, Aβ (A+) and tau-PET positivity (T+).

#### Performance of VeraBIND Tau to predict tau-PET status in A/T groups

We first assessed how VeraBIND Tau could predict tau-PET status by focusing on the individuals with concordant PET results, either supporting or excluding AD tau pathology. Specifically, we calculated the sensitivity and overall accuracy of VeraBIND Tau in identifying T+ individuals in these two groups, irrespective of cognitive status, and separately in CU and CI participants. We next evaluated the proportions of VeraBIND Tau positivity in individuals with discordant PET results (A-T+ and A+T-groups) and compared them using a Fisher’s Exact test.

#### Performance of VeraBIND Tau to predict amyloid and tau status, compared to other plasma biomarkers

The diagnostic performance of plasma VeraBIND Tau, Aβ42/40 ratio, pTau217, pTau181 and pTau231 levels to predict Aβ and tau-PET status in the entire sample was compared using receiver operating characteristic (ROC) curve analyses and DeLong tests.

We then computed the sensitivity, specificity, overall accuracy, as well as their 95% confidence intervals, for VeraBIND Tau and plasma pTau217 (i.e., the two best performing plasma biomarkers for predicting tau-PET status), using different thresholds to define positivity on the latter. We used the 0.142, 0.193, and 0.256 pg/mL cutoffs, which were demonstrated to provide 95% sensitivity, optimize sensitivity and specificity (∼92% sensitivity/specificity) and provide 95% specificity to predict amyloid-PET status, respectively, in a large multicentric cohort of 411 individuals (partially overlapping with the current dataset).^22^

Positive and negative predictive values (PPV and NPV, respectively) were calculated for the entire sample based on the observed prevalence of tau-PET positivity within the current sample of 145 participants (47.6%). To provide more accurate estimates of tau-PET positivity prevalence within each clinical group, we used the tau-PET positivity prevalence observed in a large, recently published pooled cohort analysis that included 6514 individuals, comprising CU individuals and patients with a clinical diagnosis of Mild Cognitive Impairment, AD dementia or other neurodegenerative disorders to compute PPV and NPV for each clinical group (i.e., 60% prevalence in CI individuals, 10% prevalence in CU individuals).^6^

Moreover, we compared the performance of VeraBIND Tau to plasma pTau217 in predicting tau-PET positivity in low (Braak-like stages 1-3) and high (Braak-like stages 4-6) tau pathology stages, to test their performance at early tau pathology stages. In a regression model adjusted for age, sex and education, and corrected for multiple comparisons using the Bonferroni method, we first compared the respective values of both plasma assays for each two-by-two comparison among the Braak-like 0, 1-3, and 4-6 groups. Their sensitivity and area under the curve (AUC) to predict tau-PET positivity in Braak-like 1-3 versus Braak-like 0 individuals, and in Braak-like 4-6 versus Braak-like 0 participants were compared using McNemar tests and DeLong tests, respectively. In linear regression models, we also assessed the associations between both assays and the entorhinal tau-PET signal in Braak-like 0-3 individuals, adjusting for age, sex, and education.

#### Two-test approach combining low threshold pTau217 and VeraBIND Tau to reflect the A/T stratification

As plasma pTau217 best predicted the Aý status (A) and VeraBIND Tau best predicted the tau-PET status (T), we stratified participants into four plasma A/T-like groups using the ≥0.142 pg/mL threshold for plasma pTau217 positivity (cutoff providing 95% sensitivity to predict amyloid-PET positivity)^22^ and the ≥1.0 cutoff for VeraBIND Tau positivity. We then descriptively examined how this plasma A/T-like stratification corresponded to the A/T stratification established based on the available CSF and/or PET biomarkers (i.e., A+ if Centiloid value≥20 or CSF Aβ42 levels≤544 pg/mL; T+ if Braak-like tau-PET stage>0).

#### Correlational analyses

Age-adjusted Spearman’s rank correlation coefficients were calculated between the VeraBIND Tau RLU ratio and the MMSE score, the episodic memory composite score, the entorhinal and inferior temporal tau-PET signal, the plasma concentrations of pTau217, pTau181 and pTau231, and the Centiloid value, to assess its interest as a semi-quantitative measure. Correlations were examined in the entire sample and in analyses restricted to the CU individuals.

#### Longitudinal analyses

Tau-PET data were not available longitudinally. However, a subsample of 99 individuals (64 CU, 35 CI; 48 A-T-, 10 A+T-, 38 A+T+, 3 A-T+) had two to four available MMSE scores (Median=3.0, Q1-Q3=2.0-3.0), while 83 participants (64 CU, 19 CI; 48 A-T-, 10 A+T-, 22 A+T+, 3 A-T+) had two to four available episodic memory z-scores (Median=3.0, Q1-Q3=2.0-3.0). The mean cognitive follow-up duration was of 2.6±1.0 years for both measures (min=0.53 – max=5.3). Linear mixed-effects (LME) models were fitted to examine the association between baseline VeraBIND Tau scores and longitudinal changes in MMSE and episodic memory z-scores. The models included fixed effects for time, baseline VeraBIND Tau, and their interaction, as well as random intercepts, and were adjusted for age, sex, and education. To evaluate whether these associations were independent of plasma p-Tau217, the analyses were repeated with additional adjustment for plasma p-Tau217 concentration.

Moreover, a subsample of 88 individuals (58 CU and 30 CI; 46 A-T-, 7 A+T-, 32 A+T+, and 3 A-T+) had available two to five available longitudinal VeraBIND Tau values (Median=3.0, Q1-Q3=2.0-3.0; total number of available blood samples=207). The mean follow-up duration was of 1.72±0.94 years (min=0.35 – max=3.85). We computed the annual rate of change on VeraBIND Tau for each participant using linear regression. We then examined correlations between this annual rate of change and cross-sectional entorhinal and inferior temporal tau-PET SUVr, episodic memory z-score, MMSE, plasma concentrations of pTau217, pTau181, and pTau231, using Spearman’s rank coefficients. In addition, we examined the biological and cognitive profiles of individuals who converted on VeraBIND Tau over their follow-up.

## Results

### Participants’ characteristics

Seventy-nine participants (54.5%) were classified as CU and 66 participants (45.5%) as CI (Table 1), having either mild cognitive impairment (*n*=43/66, 29.6%) or dementia (*n*=23/66, 15.9%). CU individuals were marginally younger and more educated than CI patients. The number of A+ individuals was higher in CI (*n*=52/66, 78.8%) than in CU (*n*=21/79, 26.6%) participants. Similarly, there were more T+ individuals in the CI group (*n*=52/66, 78.8%, including 47/66 A+T+) than in the CU group (*n*=17/79, 21.5%, including 13/17 A+T+). In addition, the CI group included more A+T+ individuals and less A-T-individuals than the CU group (71.2% vs. 16.5%, and 13.6% vs. 68.3%, respectively), while proportions of A-T+ and A+T-individuals did not differ between groups (7.6% in CI vs. 5.1% in CU, and 7.6% in CI vs. 10.1% in CU, respectively). As expected, MMSE and cognitive measures were lower in CI than in CU individuals, while plasma pTau species concentrations and tau-PET signal in the entorhinal cortex and inferior temporal neocortex were higher in CI than in CU individuals.

**Table 1.**
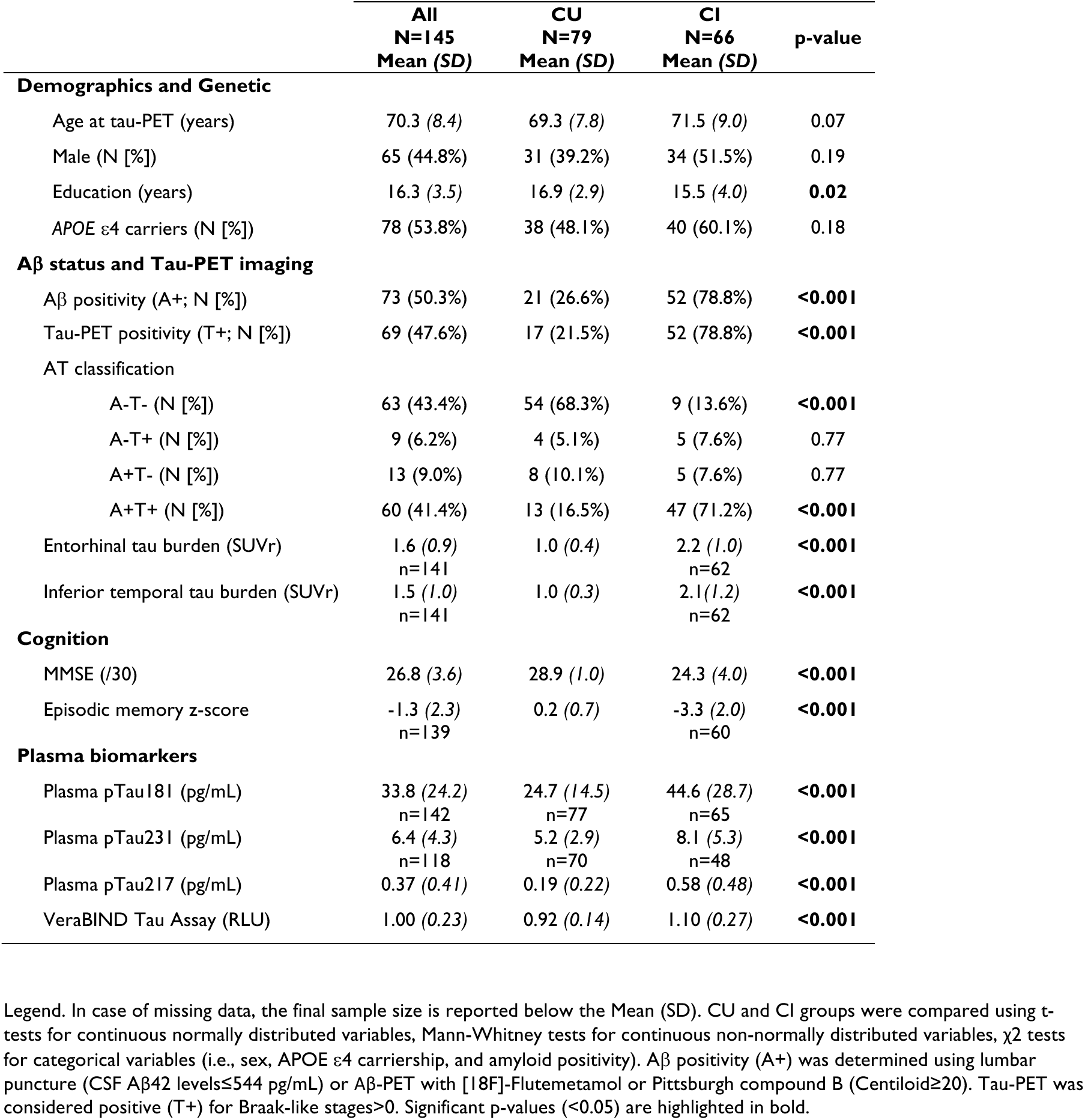
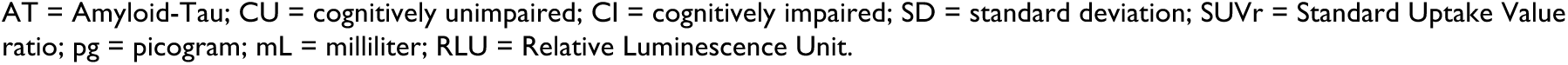
Participants’ characteristics.

|  | **All**<br>**N=145**<br>**Mean (SD)** | **CU**<br>**N=79**<br>**Mean (SD)** | **CI**<br>**N=66**<br>**Mean (SD)** | **p-value** |
| --- | --- | --- | --- | --- |
| **Demographics and Genetic** |  |  |  |  |
| Age at tau-PET (years) | 70.3 (8.4) | 69.3 (7.8) | 71.5 (9.0) | 0.07 |
| Male (N [%]) | 65 (44.8%) | 31 (39.2%) | 34 (51.5%) | 0.19 |
| Education (years) | 16.3 (3.5) | 16.9 (2.9) | 15.5 (4.0) | **0.02** |
| APOE ε4 carriers (N [%]) | 78 (53.8%) | 38 (48.1%) | 40 (60.1%) | 0.18 |
| **Aβ status and Tau-PET imaging** |  |  |  |  |
| Aβ positivity (A+; N [%]) | 73 (50.3%) | 21 (26.6%) | 52 (78.8%) | **<0.001** |
| Tau-PET positivity (T+; N [%]) | 69 (47.6%) | 17 (21.5%) | 52 (78.8%) | **<0.001** |
| AT classification |  |  |  |  |
| A-T- (N [%]) | 63 (43.4%) | 54 (68.3%) | 9 (13.6%) | **<0.001** |
| A-T+ (N [%]) | 9 (6.2%) | 4 (5.1%) | 5 (7.6%) | 0.77 |
| A+T- (N [%]) | 13 (9.0%) | 8 (10.1%) | 5 (7.6%) | 0.77 |
| A+T+ (N [%]) | 60 (41.4%) | 13 (16.5%) | 47 (71.2%) | **<0.001** |
| Entorhinal tau burden (SUVR) | 1.6 (0.9)<br>n=141 | 1.0 (0.4) | 2.2 (1.0)<br>n=62 | **<0.001** |
| Inferior temporal tau burden (SUVR) | 1.5 (1.0)<br>n=141 | 1.0 (0.3) | 2.1 (1.2)<br>n=62 | **<0.001** |
| **Cognition** |  |  |  |  |
| MMSE (/30) | 26.8 (3.6) | 28.9 (1.0) | 24.3 (4.0) | **<0.001** |
| Episodic memory z-score | -1.3 (2.3)<br>n=139 | 0.2 (0.7) | -3.3 (2.0)<br>n=60 | **<0.001** |
| **Plasma biomarkers** |  |  |  |  |
| Plasma pTau181 (pg/mL) | 33.8 (24.2)<br>n=142 | 24.7 (14.5)<br>n=77 | 44.6 (28.7)<br>n=65 | **<0.001** |
| Plasma pTau231 (pg/mL) | 6.4 (4.3)<br>n=118 | 5.2 (2.9)<br>n=70 | 8.1 (5.3)<br>n=48 | **<0.001** |
| Plasma pTau217 (pg/mL) | 0.37 (0.41) | 0.19 (0.22) | 0.58 (0.48) | **<0.001** |
| VeraBIND Tau Assay (RLU) | 1.00 (0.23) | 0.92 (0.14) | 1.10 (0.27) | **<0.001** |

Legend. In case of missing data, the final sample size is reported below the Mean (SD). CU and CI groups were compared using t-tests for continuous normally distributed variables, Mann-Whitney tests for continuous non-normally distributed variables, $\chi^2$ tests for categorical variables (i.e., sex, APOE ε4 carriership, and amyloid positivity). Aβ positivity (A+) was determined using lumbar puncture (CSF Aβ42 levels ≤ 544 pg/mL) or Aβ -PET with [18F]-Flutemetamol or Pittsburgh compound B (Centiloid ≥ 20). Tau-PET was considered positive (T+) for Braak-like stages $>0$ . Significant p-values ( $<0.05$ ) are highlighted in bold.
AT = Amyloid-Tau; CU = cognitively unimpaired; CI = cognitively impaired; SD = standard deviation; SUVr = Standard Uptake Value ratio; pg = picogram; mL = milliliter; RLU = Relative Luminescence Unit.

### VeraBIND Tau result is highly consistent with tau-PET status

In individuals with consistent PET results (i.e., A+T+, A-T-; Table 2), VeraBIND Tau was positive in 3.2% of the A-T-individuals (*n*=2/63, including one CU and one CI), and 96.7% of the A+T+ individuals (*n*=58/60, two CI patients having a negative result), achieving an overall accuracy of 96.7%. The diagnostic accuracy was relatively similar across clinical groups (CU: *n*=66/67, 98.5%; CI: *n*=53/56, 94.6%). The only A-T-CI patient with positive VeraBIND Tau result had apraxia of speech, a clinical phenotype suspected to be due to corticobasal degeneration, a primary 4R-tauopathy that is not detected by [^18^F]MK6240 tau-PET.^59,60^

**Table 2.** Results of VeraBIND Tau Test by clinical and A/T status.

|  | A-T- |  | A+T- |  | A+T+ |  | A-T+ |  |
| --- | --- | --- | --- | --- | --- | --- | --- | --- |
| CU (n=79) |  |  |  |  |  |  |  |  |
| VeraBIND Tau result (N -; N +) | 53 - | 1 + | 8 - | 0 + | 0 - | 13 + | 2 - | 2 + |
| VeraBIND Tau positivity rate (%) | 1.9% |  | 0.0% |  | 100% |  | 50.0% |  |
| CI (n=66) |  |  |  |  |  |  |  |  |
| VeraBIND Tau result (N -; N +) | 8 - | 1 + | 4 - | 1 + | 2 - | 45 + | 0 - | 5 + |
| VeraBIND Tau positivity rate (%) | 11.1% |  | 20.0% |  | 95.7% |  | 100.0% |  |
| Entire sample (n=145) |  |  |  |  |  |  |  |  |
| VeraBIND Tau result (N -; N +) | 61 - | 2 + | 12 - | 1 + | 2 - | 58 + | 2 - | 7 + |
| VeraBIND Tau positivity rate (%) | 3.2% |  | 7.7% |  | 96.7% |  | 77.8% |  |

Legend. CU = cognitively unimpaired, CI = cognitively impaired. The - and + symbols denote the negative or positive test result. N - = number of individuals with a negative VeraBIND Tau result; N + = number of individuals with a positive VeraBIND Tau result. The Aβ (A) status was determined using lumbar puncture (positive for CSF Aβ42 levels ≤ 544 pg/mL) or Aβ -PET (positive for Centiloid ≥ 20). The tau (T) status was defined using [18F]MK6240 tau-PET (positive for Braak-like tau-PET stages $>$ 0). The A-T- individual with a positive VeraBIND Tau result has a clinical diagnosis of apraxia of speech, a clinical phenotype suspected of cortico-basal degeneration.

In individuals with discordant PET results (i.e., A+T-, A-T+; Table 2), VeraBIND Tau was negative in all but one A+T-participants (*n*=1/13, 7.7%), who was CI. Moreover, positive VeraBIND Tau results were observed in 77.8% (*n*=7/9) of the A-T+ cases, including all five A-T+ CI patients (Braak-like stages 1-2 in four cases and one case with Braak-like stage 4 and subthreshold amyloidosis), suspected to suffer from primary age-related tauopathy (PART, *n*=4) or fronto-temporal lobar degeneration (FTLD, *n*=1), and two A-T+ CU individuals who were assigned a Braak-like stage 3. The two A-T+ cases with a negative VeraBIND Tau score were CU and were assigned a Braak-like stage of 1 or 2. The proportion of positive VeraBIND Tau results was higher in A-T+ cases (n=7/9, 77.8%) than in A+T-cases (*n*=1/13, 7.7%; Fisher’s Exact test *p*=0.001), indicating greater concordance between VeraBIND Tau and tau-PET status than with Aβ status.

### Comparison of VeraBIND Tau with other plasma biomarkers

ROC curves predicting tau-PET status computed on the entire sample (*n*=145; Fig. 2.A) indicated that the largest AUC was found for VeraBIND Tau (AUC=0.97), followed by pTau217 plasma concentration (AUC=0.92). DeLong Tests indicated that VeraBIND Tau better predicted tau-PET positivity compared to all the other blood-based biomarkers (VeraBIND Tau vs. pTau217: *z*=1.96, *p*=0.049; VeraBIND Tau vs. pTau181 [missing data=3], pTau231 [missing data=27] or Aβ42/Aβ40 [missing data=6]: all *z*’s>3.68, *p*<0.001), while the highest AUC (AUC=0.93) for predicting the Aβ status was found for plasma pTau217 (Fig. 2.B; DeLong Tests: pTau217 vs. VeraBIND Tau: *z*=1.89, *p*=0.058; pTau217 vs. pTau181: *z*=3.72, *p*=0.0002; pTau217 vs. pTau231: *z*=4.37, *p*<0.001; pTau217 vs. Aβ42/Aβ40: *z*=3.03, *p*=0.002).

**Fig. 2.**
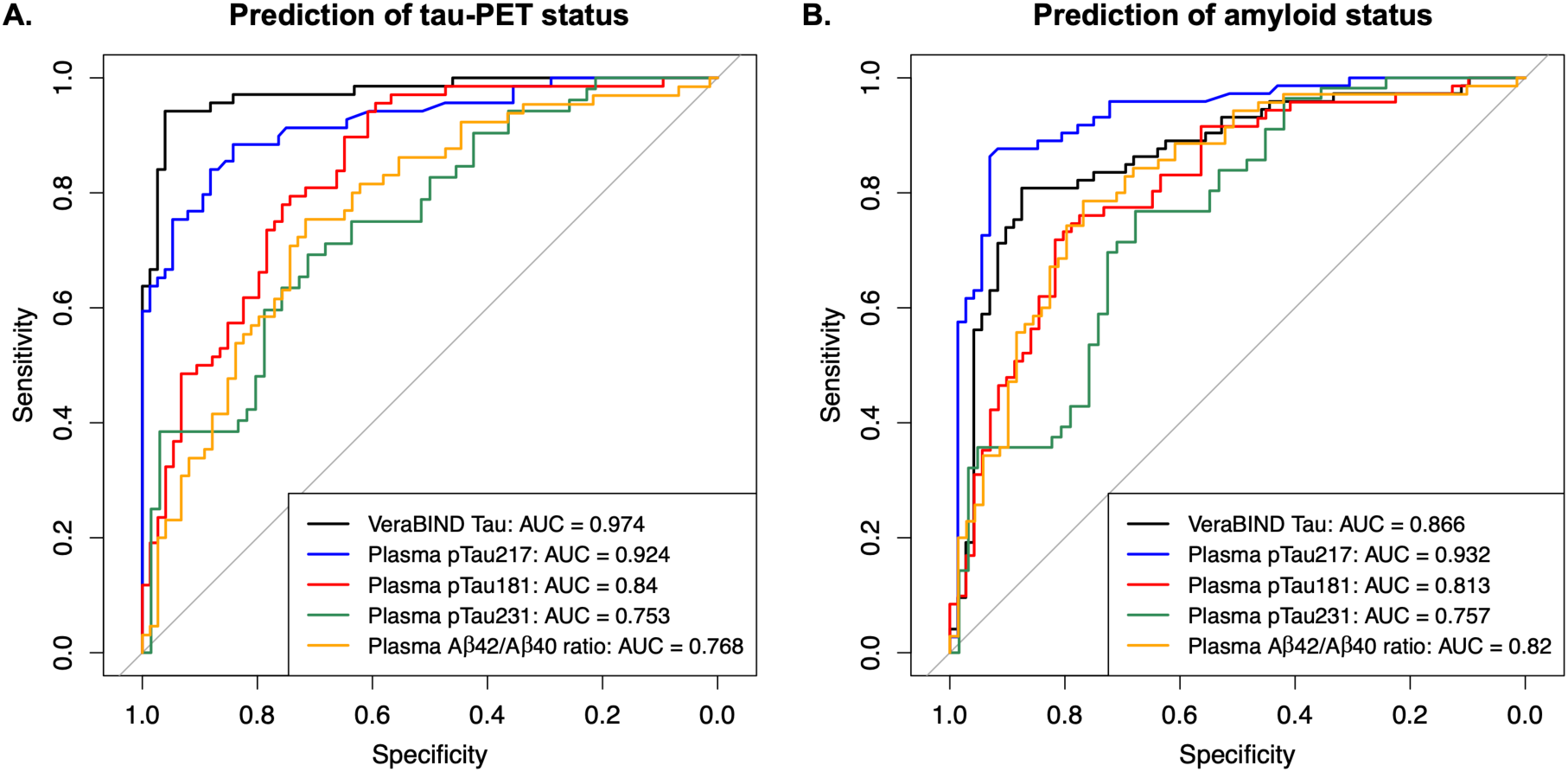
Receiver Operating Characteristic (ROC) curves for predicting tau and amyloid positivity. ROC curves compare the performance of VeraBIND Tau, plasma pTau217, pTau181 (missing data=3), pTau231 (missing data=27), and the Aý42/Aý40 ratio (missing data=6) for predicting: (**A).** tau-PET positivity (Braak-like tau-PET stage>0), and (**B).** amyloid positivity (Centiloid≥20 or CSF Aβ42≤544 pg/mL).

The sensitivity, specificity, overall accuracy, PPV and NPV for tau-PET positivity were systematically higher for VeraBIND Tau than for plasma pTau217 (the second-best performing plasma biomarker, Table 3), regardless of the threshold used. Remarkably, the VeraBIND Tau PPV was 85.9% in CU participants, while the highest PPV found for pTau217 only achieved 57.5% at the 0.256pg/mL threshold.

**Table 3.**
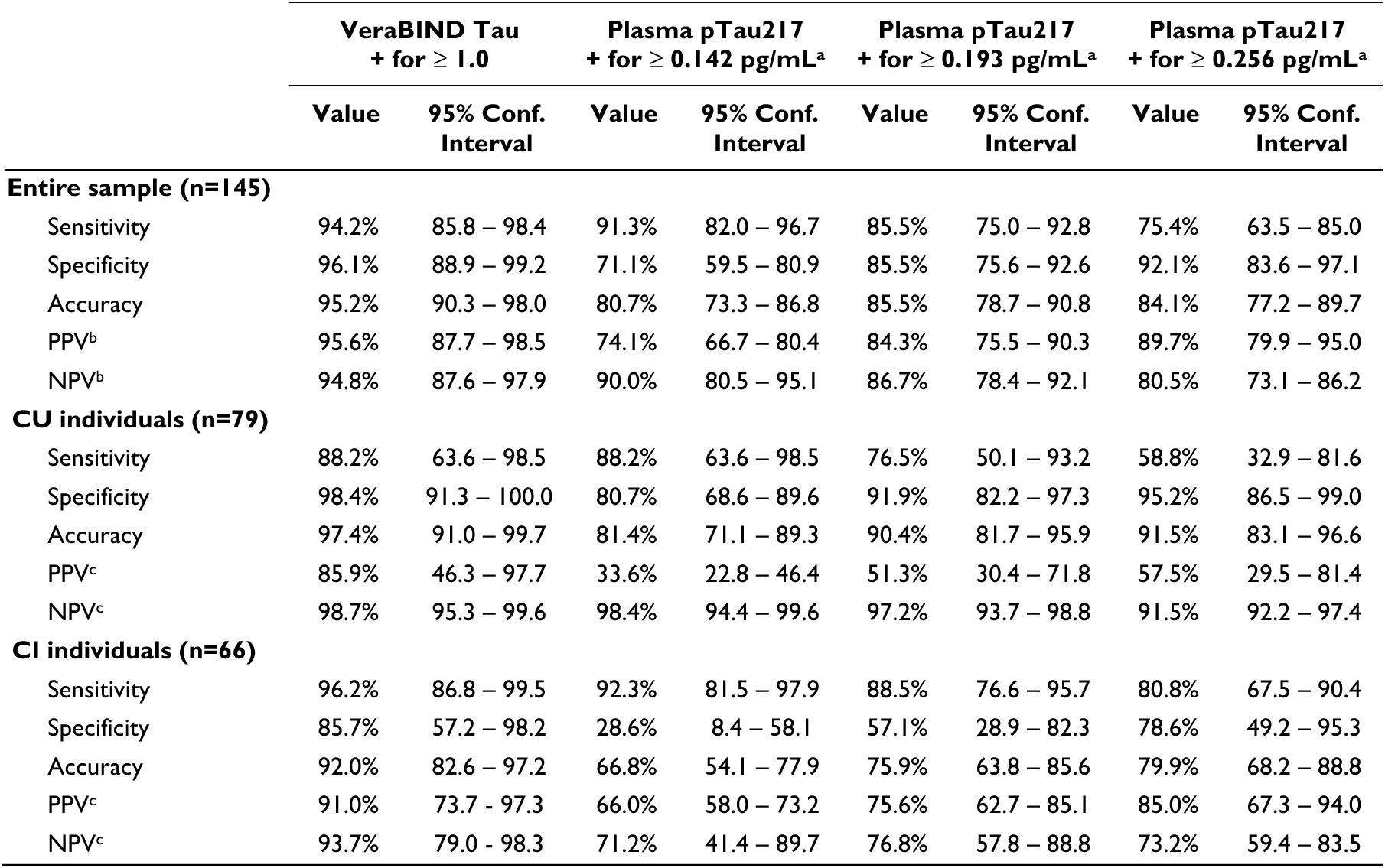
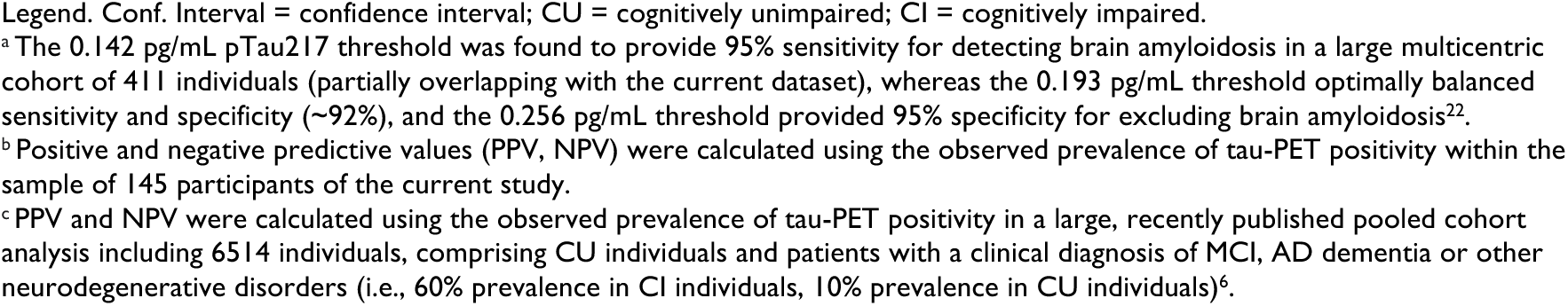
Diagnostic performance of VeraBIND Tau Test for predicting tau-PET positivity (Braak-like tau-PET stage>0), compared to plasma pTau217.

|  | **VeraBIND Tau**<br>**+ for ≥ 1.0** |  | **Plasma pTau217**<br>**+ for ≥ 0.142 pg/mL<sup>a</sup>** |  | **Plasma pTau217**<br>**+ for ≥ 0.193 pg/mL<sup>a</sup>** |  | **Plasma pTau217**<br>**+ for ≥ 0.256 pg/mL<sup>a</sup>** |  |
| --- | --- | --- | --- | --- | --- | --- | --- | --- |
|  | **Value** | **95% Conf.**<br>**Interval** | **Value** | **95% Conf.**<br>**Interval** | **Value** | **95% Conf.**<br>**Interval** | **Value** | **95% Conf.**<br>**Interval** |
| **Entire sample (n=145)** |  |  |  |  |  |  |  |  |
| Sensitivity | 94.2% | 85.8 – 98.4 | 91.3% | 82.0 – 96.7 | 85.5% | 75.0 – 92.8 | 75.4% | 63.5 – 85.0 |
| Specificity | 96.1% | 88.9 – 99.2 | 71.1% | 59.5 – 80.9 | 85.5% | 75.6 – 92.6 | 92.1% | 83.6 – 97.1 |
| Accuracy | 95.2% | 90.3 – 98.0 | 80.7% | 73.3 – 86.8 | 85.5% | 78.7 – 90.8 | 84.1% | 77.2 – 89.7 |
| PPV<sup>b</sup> | 95.6% | 87.7 – 98.5 | 74.1% | 66.7 – 80.4 | 84.3% | 75.5 – 90.3 | 89.7% | 79.9 – 95.0 |
| NPV<sup>b</sup> | 94.8% | 87.6 – 97.9 | 90.0% | 80.5 – 95.1 | 86.7% | 78.4 – 92.1 | 80.5% | 73.1 – 86.2 |
| **CU individuals (n=79)** |  |  |  |  |  |  |  |  |
| Sensitivity | 88.2% | 63.6 – 98.5 | 88.2% | 63.6 – 98.5 | 76.5% | 50.1 – 93.2 | 58.8% | 32.9 – 81.6 |
| Specificity | 98.4% | 91.3 – 100.0 | 80.7% | 68.6 – 89.6 | 91.9% | 82.2 – 97.3 | 95.2% | 86.5 – 99.0 |
| Accuracy | 97.4% | 91.0 – 99.7 | 81.4% | 71.1 – 89.3 | 90.4% | 81.7 – 95.9 | 91.5% | 83.1 – 96.6 |
| PPV<sup>c</sup> | 85.9% | 46.3 – 97.7 | 33.6% | 22.8 – 46.4 | 51.3% | 30.4 – 71.8 | 57.5% | 29.5 – 81.4 |
| NPV<sup>c</sup> | 98.7% | 95.3 – 99.6 | 98.4% | 94.4 – 99.6 | 97.2% | 93.7 – 98.8 | 91.5% | 92.2 – 97.4 |
| **CI individuals (n=66)** |  |  |  |  |  |  |  |  |
| Sensitivity | 96.2% | 86.8 – 99.5 | 92.3% | 81.5 – 97.9 | 88.5% | 76.6 – 95.7 | 80.8% | 67.5 – 90.4 |
| Specificity | 85.7% | 57.2 – 98.2 | 28.6% | 8.4 – 58.1 | 57.1% | 28.9 – 82.3 | 78.6% | 49.2 – 95.3 |
| Accuracy | 92.0% | 82.6 – 97.2 | 66.8% | 54.1 – 77.9 | 75.9% | 63.8 – 85.6 | 79.9% | 68.2 – 88.8 |
| PPV<sup>c</sup> | 91.0% | 73.7 – 97.3 | 66.0% | 58.0 – 73.2 | 75.6% | 62.7 – 85.1 | 85.0% | 67.3 – 94.0 |
| NPV<sup>c</sup> | 93.7% | 79.0 – 98.3 | 71.2% | 41.4 – 89.7 | 76.8% | 57.8 – 88.8 | 73.2% | 59.4 – 83.5 |

Legend. Conf. Interval = confidence interval; CU = cognitively unimpaired; CI = cognitively impaired.
<sup>a</sup> The 0.142 pg/mL pTau217 threshold was found to provide 95% sensitivity for detecting brain amyloidosis in a large multicentric cohort of 411 individuals (partially overlapping with the current dataset), whereas the 0.193 pg/mL threshold optimally balanced sensitivity and specificity (~92%), and the 0.256 pg/mL threshold provided 95% specificity for excluding brain amyloidosis<sup>22</sup>.
<sup>b</sup> Positive and negative predictive values (PPV, NPV) were calculated using the observed prevalence of tau-PET positivity within the sample of 145 participants of the current study.
<sup>c</sup> PPV and NPV were calculated using the observed prevalence of tau-PET positivity in a large, recently published pooled cohort analysis including 6514 individuals, comprising CU individuals and patients with a clinical diagnosis of MCI, AD dementia or other neurodegenerative disorders (i.e., 60% prevalence in CI individuals, 10% prevalence in CU individuals)<sup>6</sup>.

### VeraBIND Tau outperforms plasma pTau217 to detect low tau pathology stages

To identify the participants driving the additional accuracy of VeraBIND Tau vs. pTau217 plasma concentration, we compared diagnostic performances of both plasma biomarkers in low (Braak-like 1-3) and high tau-PET stages (Braak-like 4-6; Fig. 3.A-B). In a regression model adjusted for age, sex and education, and corrected for multiple comparisons using the Bonferroni method, the VeraBIND Tau Score was elevated in both Braak-like 1-3 (*Mean*=1.10, *SE*=0.05, β=0.24, 95% confidence interval [0.11-0.36], *p*<0.001) and Braak-like 4-6 groups (*Mean*=1.16, *SE*=0.03, β=0.29, 95% confidence interval [0.20-0.37], *p*<0.001), compared to Braak-like 0 individuals (*Mean*=0.87, *SE*=0.02), while Braak-like 1-3 and Braak-like 4-6 groups did not differ (β=-0.05, 95% confidence interval [-0.07-0.18], *p*=0.98). In contrast, plasma pTau217 was increased in the Braak-like 4-6 group (*Mean*=0.71, *SE*=0.04), compared to both Braak-like 0 (*Mean*=0.134, *SE*=0.04, β=0.58, 95% confidence interval [0.44-0.72], *p*<0.001) and 1-3 groups (*Mean*=0.320, *SE*=0.08, β=0.40, 95% confidence interval [0.18-0.61], *p*<0.001), which did not differ (β=0.19, 95% confidence interval [-0.02-0.40], *p*=0.10).

**Fig. 3.**
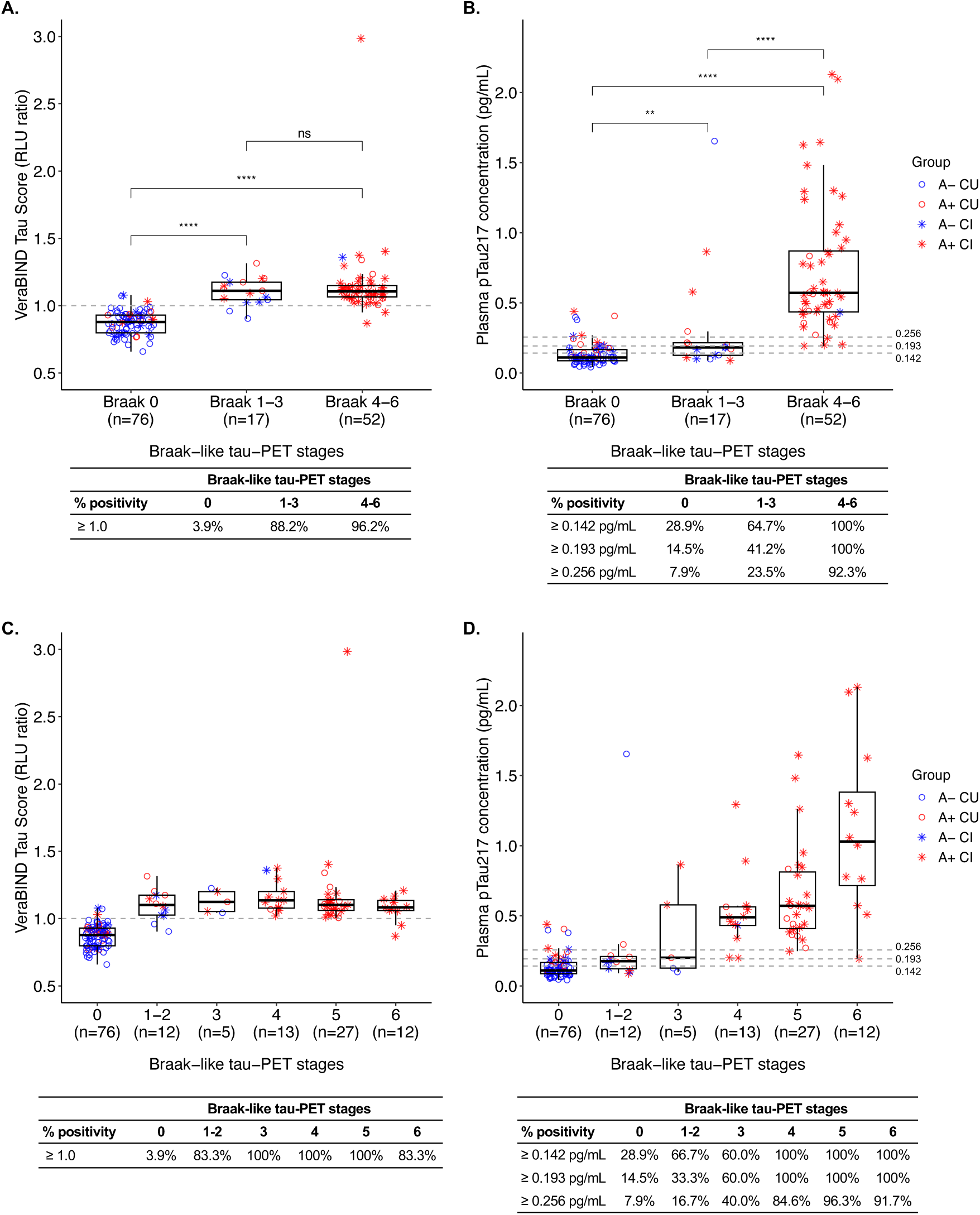
Plasma levels and positivity rates of VeraBIND Tau and plasma pTau217 across Braak-like tau-PET stage groups. **(A–B)** Participants were classified into three Braak-like tau-PET stage groups: Braak-like 0 (visually tau-PET negative), low (Braak-like stages 1–3), and advanced (Braak-like stages 4–6). **(C–D)** Plasma levels and positivity rates of VeraBIND Tau and plasma pTau217 are shown for each Braak-like tau-PET stage group. Boxplots show the median and interquartile range (IQR; 25th–75th percentile), with whiskers extending to 1.5 × IQR. Jittered points represent individual observations. VeraBIND Tau Score (RLU ratio) is considered positive for Scores ≥ 1.0. For plasma pTau217, we used the 0.142, 0.193, and 0.256 pg/mL cutoffs, which were demonstrated to provide 95% sensitivity, optimize sensitivity and specificity (∼92% sensitivity/specificity) and provide 95% specificity to predict amyloid-PET status, respectively, in a large multicentric cohort.^22^ pg = picogram; mL = milliliter; A-= amyloid negative; A+ = amyloid positive; CU = cognitively unimpaired; CI = cognitively impaired. ns *p* ≥ .05; ** *p* < .01, **** *p <* .0001

In Braak-like 1-3 individuals, VeraBIND Tau was positive in 88.2% (*n*=15/17) individuals, whereas pTau217 was positive in 64.7% (*n*=11/17), 41.2% (*n*=7/11), and 23.5% (*n*=4/17) individuals at the 0.142, 0.193, and 0.256 thresholds, respectively (Fig. 3.A-B). McNemar tests indicated that VeraBIND Tau sensitivity was higher than pTau217 sensitivity at the 0.193 (*χ*^2^=4.90, *p*=0.03) and 0.256 cutoffs (χ^2^=7.69, *p*=0.006). At the 0.142 threshold, the sensitivity did not differ between VeraBIND Tau and pTau217 (χ^2^=1.1, *p*=0.29), but at this cutoff, pTau217 was also positive in 28.9% (*n*=22/76) Braak-like 0 participants (T-) against only 3.9% (*n*=3/76) for VeraBIND Tau. The AUC for detecting tau-PET signals in Braak-like 1-3 vs. Braak-like 0 individuals was higher for VeraBIND Tau (AUC=0.96) than for plasma pTau217 concentration (AUC=0.74, *z*=2.99, *p*=0.003). In contrast, VeraBIND Tau (*n*=50/52, 96.2% sensitivity, AUC=0.98) and pTau217 (0.142 and 0.193 thresholds: *n*=52/52, 100% sensitivity; 0.256 threshold: *n*=48/52, 92.3% sensitivity; AUC=0.98) had similar sensitivity and AUC (*z*=-0.43, *p*=0.66) to distinguish individuals with Braak-like 4-6 from Braak-like 0 participants. Importantly, VeraBIND Tau demonstrated high sensitivity for detecting tau-PET positivity even at the earliest Braak-like stages 1-2, whereas plasma p-tau217 exhibited broad dynamic range across Braak stages (Fig. 3.C-D). In a regression model adjusted for age, sex, and education, and limited to 93 individuals with Braak-like stage 0–3, VeraBIND Tau demonstrated a significant association with entorhinal tau-PET signal (β=1.69, 95% confidence interval [1.27-2.10], *p*<0.001), whereas plasma pTau217 concentration did not (β=0.31, 95% confidence interval [-0.06-0.68], *p*=0.10).

### A two-test approach combining plasma pTau217 and VeraBIND Tau accurately diagnoses presymptomatic AD

The concordance rate between the plasma A/T-like stratification (i.e., positive plasma pTau217 if ≥0.142 pg/mL; positive VeraBIND Tau score if ≥ 1.0) and the AT stratification established on CSF and/or PET biomarkers (i.e., A+ if Centiloid value≥20 or CSF Aβ42 levels≤544 pg/mL; T+ if Braak-like tau-PET stage>0) was 83.5% in CU individuals (Fig. 4.A), 78.8% in CI patients (Fig. 4.B), 81.5% in the entire sample (Fig. 4.C.). All negative CI patients on both plasma biomarkers were A-T-when assessed using CSF and/or PET. Importantly, this two-test approach diagnosed 100% of A+T+ CU individuals, as both plasma biomarkers were positive in all A+T+ CU (*n*=13/13). Moreover, no other CU individual (i.e., A-T-, A+T-, A-T+) had both positive pTau217 and VeraBIND Tau results (i.e., they were all non pTau217+ VBT+). In the entire sample, both plasma biomarkers were positive in 93.3% (*n*=56/60) of A+T+ individuals, while 95.3% (*n*=81/85) of the other individuals (i.e., non A+T+ including A-T-, A+T-, A-T+) were not positive on both plasma biomarkers. Overall, this two-test plasma approach accurately identified A+T+ individuals compared with all other biomarker profiles in both CU and CI individuals.

**Fig. 4.**
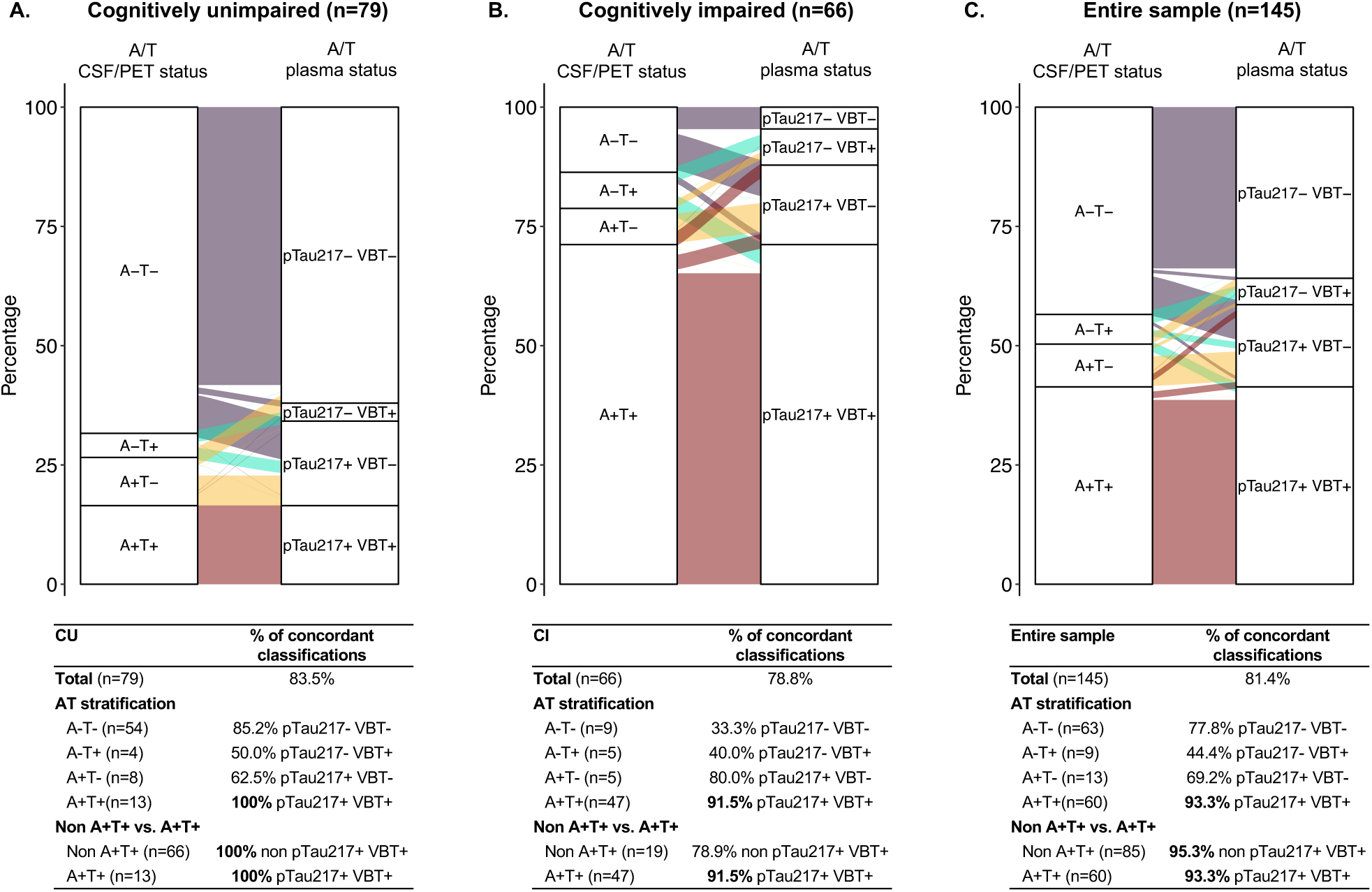
Alluvial plots illustrating the concordance between the amyloid/tau (A/T) CSF/PET stratification and the plasma Lumipulse pTau217/VeraBIND Tau stratification in : (**A)** Clinically unimpaired individuals (CU, n=79), (**B)** Clinically impaired participants (CI, n=66), and (**C)** The entire sample (n=145). CSF = cerebro-spinal fluid; PET = positron emission tomography; VBT = VeraBIND Tau assay. The – and + symbols denote the negative or positive test results. The A status for the CSF/PET stratification was positive for Centiloid values ≥20 or CSF Aβ42 levels ≤544 pg/mL, whereas the T status was positive for Braak-like tau-PET stages >0. The plasma pTau217 status was positive (pTau217+) for concentrations ≥0.142 pg/mL. This cutoff demonstrated 95% sensitivity to predict amyloid-PET status in a large multicentric cohort of 411 individuals (partially overlapping with the current dataset).^22^ The VeraBIND Tau status was positive (VBT+) for relative radioluminescence unit (RLU) ratio scores ≥1.0. The non A+T+ group comprised A-T-, A-T+, and A+T+ individuals.

### VeraBIND Tau semi-quantitative measure is associated with cognitive, tau-PET and plasma measures

To assess the utility of the VeraBIND Tau score as a semi-quantitative measure, we correlated this novel plasma measure with the MMSE (Fig. 5.A), an episodic memory composite score (Fig. 5.B), the entorhinal and inferior temporal tau-PET signal (SUVr; Fig. 5.C-D), the plasma concentration of pTau217 (Fig. 5.E), pTau181 (Fig. 5.F; missing data=3), and pTau231 (missing data=27), as well as the Centiloid value (missing data=49; Supplementary Fig. 1). All these cross-sectional associations were significant (all age-adjusted Spearman’s rho>0.35, all p-values<0.001). Consistent with ROC curve analysis, in the subsample of 96 participants with both available Centiloid and regional tau-PET data (74 CU, 22 CI; 57 A-T-, 9 A+T-, 23 A+T+, 7 A-T+), the VeraBIND Tau score was more strongly associated with the entorhinal tau-PET signal (age-adjusted *r_s_*=0.63, *p*<0.001) than with the Centiloid value (age-adjusted *r_s_*=0.40, *p*<0.001; *Λ1r_s_*=-0.23, *p*=0.024). The correlation with the inferior temporal tau-PET signal was numerically higher than that with the Centiloid value (age-adjusted *r_s_*=0.52, *p*<0.001; *Λ1r_s_*=-0.12, *p*=0.30; Supplementary Fig. 1).

**Fig. 5.**
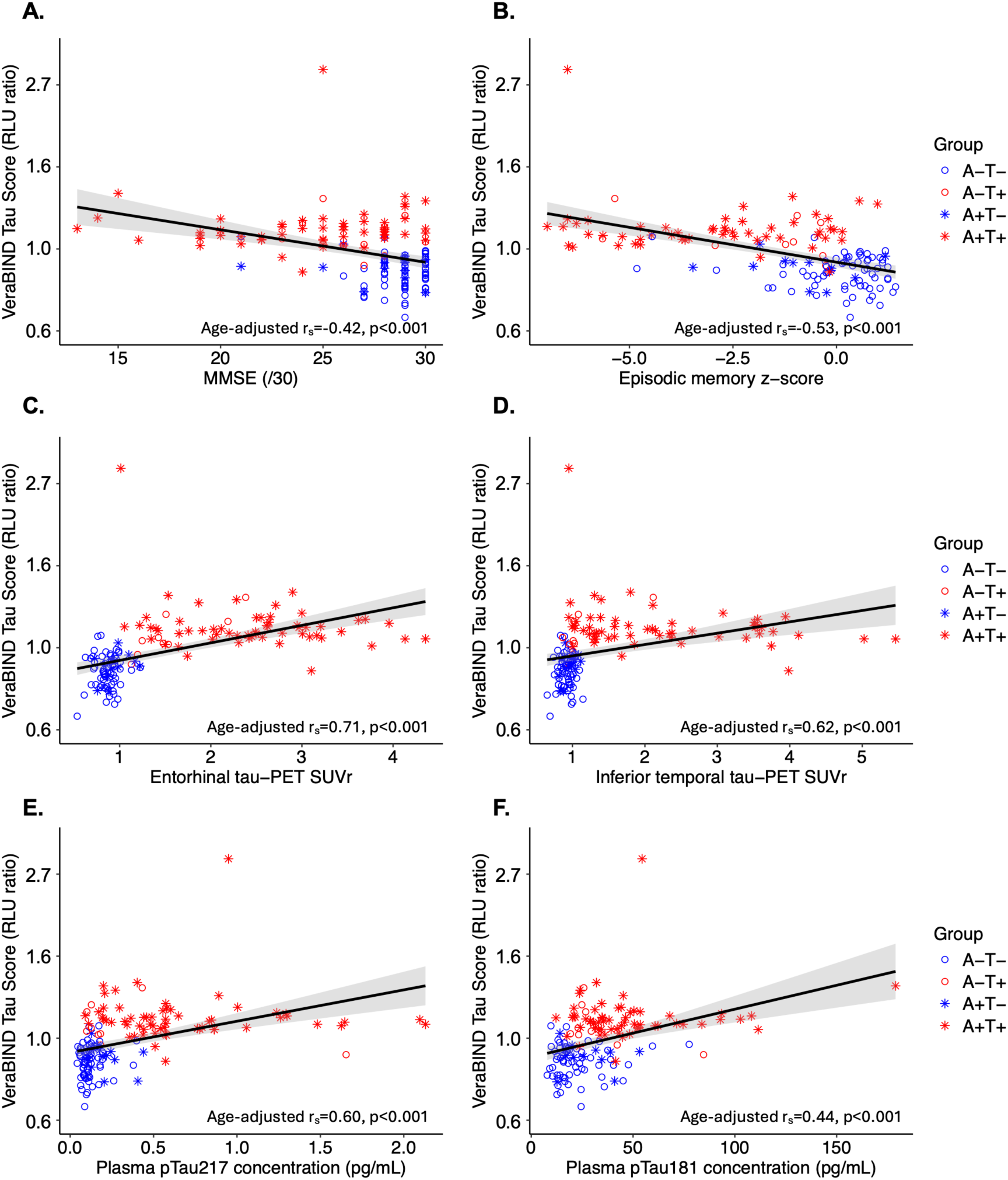
Associations of VeraBIND Tau with cognitive, regional tau-PET signal and plasma pTau biomarkers. Scatterplots showing associations between the VeraBIND Tau semi-quantitative measure (relative luminescence unit [RLU] ratio) and: (**A)** Mini-Mental State Examination score (MMSE), (**B)** Episodic memory composite score (z-score), (**C)** Entorhinal tau burden, as measured using [^18^F]MK6240 tau-PET Standard Uptake Value ratio (SUVr), **(D)** Inferior temporal tau burden (SUVr), **(E)** Plasma concentration of pTau217 (pg/mL), (**F)** Plasma concentration of pTau181 (pg/mL, missing data=3). VeraBIND Tau semi-quantitative scores were plotted on a logarithmic scale. r_s_ = Spearman’s rho; A-= amyloid negative; A+ = amyloid positive; CU = cognitively unimpaired; CI = cognitively impaired.

In CU participants only, high VeraBIND Tau scores remained associated with high entorhinal tau-PET signal (*r_s_*=0.52, *p*<0.0001), inferior temporal tau-PET signal (*r_s_*=0.43, *p*<0.0001), plasma pTau217 (*r_s_*=0.42, *p*=0.0001) and pTau181 (missing data=2; *r_s_*=0.39, *p*=0.0005), as well as the Centiloid value (missing data=5; *r_s_*=0.37, *p*=0.001), indicating that these associations were not entirely driven by the CI participants. Consistent with the relatively low variability of cognitive performance on standard neuropsychological tests in CU individuals, VeraBIND Tau scores were not associated with the MMSE scores (*r_s_*=0.10, *p*=0.38) and showed only marginal association with the episodic memory scores (*r_s_*=-0.20, *p*=0.08). A marginal association was also found with plasma pTau231 concentration in CU individuals (available in 70/79 individuals, *r_s_*=0.23, *p*=0.054).

In subsamples of participants with available cognitive follow-up, higher baseline VeraBIND Tau scores were associated with a steeper decline in MMSE (β=-3.59, *SE*=0.47, *t*=-7.63, *p*<0.001), and in episodic memory z-score (β=-0.73, *SE*=0.25, *t*=-2.89, *p*=0.004; Supplementary Table 1). These associations were unchanged after additional adjustment for plasma p-Tau217 concentration (MMSE: β=-3.58, *SE*=0.47, *t*=-7.63, *p*<0.001; episodic memory composite: β=-0.75, *SE*=0.25, *t*=-2.97, *p*=0.003).

### VeraBIND Tau annual rate of change is associated with disease severity proxy measures

The annual change in VeraBIND Tau Score correlated with cross-sectional entorhinal tau-PET SUVr (age-adjusted *r_s_* =0.23, *p*=0.03), Braak-like stages (*r_s_*=0.29, *p*=0.006), MMSE scores (*r_s_*=-0.38, *p*=0.0003), episodic memory z-score (*r_s_*=-0.33, *p*=0.002), plasma concentrations of pTau217 (*r_s_*=0.29, *p*=0.007), and pTau181 (*r_s_*=0.30, *p*=0.005), suggesting that VeraBIND Tau rate of change increases with disease progression. The association between the annual VeraBIND Tau change with pTau231 plasma concentration (available in 84/88 participants; *r_s_*=0.08, *p*=0.48) and the Centiloid value (available in 72/88 participants; *r_s_*=0.16, *p*=0.18) was not significant, while the association with the inferior temporal tau-PET SUVr was marginal (*r_s_*=0.21, *p*=0.054).

Five participants (5.7%) initially negative on VeraBIND Tau (VBT-) converted to positivity (VBT+) during follow-up (Supplementary Fig. 2), while the other participants remained either positive (*n*=32/88, 36.4%) or negative (*n*=51/88, 57.9%). Progressors included three A-T-CU individuals who were VBT- at the time of baseline tau-PET, one A+T+ CU individual with a Braak-like stage 2 who had three closely spaced datapoints and was VBT+ at the datapoint closest in time to baseline tau-PET, and one A+T+ CI participant with a Braak-like stage 4 who was VBT-two years before tau-PET but VBT+ at the datapoint closest in time to baseline tau-PET.

## Discussion

In this study, we assessed VeraBIND Tau, a new plasma biomarker designed to reflect pathological pTau binding activity, against [^18^F]MK6240 tau-PET imaging as standard of truth. Over a sample of 145 individuals, we found that the VeraBIND Tau assay outperformed the plasma levels of pTau217, pTau181, and pTau231 to predict tau-PET positivity, while, as in previous work^14,61–65^, plasma pTau217 best predicted Aβ positivity. Accordingly, the VeraBIND Tau semi-quantitative measure was more strongly associated with regional tau-PET signal than with the Centiloid value. The high diagnostic performance of VeraBIND Tau was driven by the detection of individuals with low stages of brain tau deposition (Braak-like tau-PET stages 1-3). Consequently, VeraBIND Tau provided high positive predictive value (85.9%) in CU individuals, opening the possibility of screening older adults, which is not feasible with previously available plasma tests (e.g., pTau217 PPV=57.5% using a high threshold of 0.256pg/mL). Importantly, a two-test approach combining low threshold plasma pTau217 (≥0.142pg/mL) and VeraBIND Tau identified 91.5% of A+T+ CI patients and detected 100% of A+T+ CU individuals while excluding all CU participants with other biomarker profiles.

The development of plasma biomarkers reflecting tau aggregated pathology is of utmost importance, given its high prediction of cognitive decline.^5,6^ The National Institute on Aging and the Alzheimer’s Association (NIA-AA) revised criteria for diagnosis and staging of AD^1^ positioned emerging biofluid measures of pTau205^66,67^ and microtubule-binding region containing residue 243 (MTBR-tau243)^68^ as Core 2 biomarkers, reflecting aggregated tau, besides tau-PET imaging. However, unlike tau-PET, none of these Core 2 fluid biomarkers are currently validated for clinical use. Both these fluid biomarkers have demonstrated high accuracy to predict tau-PET results when measured in the cerebrospinal fluid.^66–68^ In plasma, the endogenously cleaved MTBR-tau243 (eMTBR-tau243) showed the strongest correlation with tau-PET signal and cognition, compared to pTau205 and pTau217, in mixed samples of CI and CU individuals.^61^ In addition, the eMTBR-tau243 outperformed pTau205 and pTau217 to predict tau-PET status, especially at intermediate and advanced stages of NFT pathology (i.e., Braak-like tau-PET stages≥3). However, although its accuracy to predict abnormal tau status in low Braak-like tau-PET stages was higher than pTau205, it was only comparable to pTau217.^61^ Development of high performance biomarkers to identify individuals with advanced Braak stages continue to progress, as evidenced by another ultrasensitive plasma assay targeting N-terminal containing tau fragments.^69^ Unfortunately, much of the recent assay development results continue to suggest that tracking early tau aggregation in blood remains challenging.

The major finding of this study was that the VeraBIND Tau assay showed high diagnostic performance to detect tau-PET positivity at both low (≤ 3; AUC=0.96) and high (≥4; AUC=0.98) Braak-like stages. Specifically, detection of individuals with low Braak-like stages was better with VeraBIND Tau (AUC=0.96, 88.2% sensitivity) than with pTau217 (AUC=0.74, 64.7% sensitivity using a low threshold=0.142pg/mL). In contrast, the ability to detect individuals with high Braak-like stages was comparable for both assays (AUC=0.98). Furthermore, in individuals with Braak-like stages 0-3, the VeraBIND Tau semi-quantitative measure was associated with the entorhinal tau-PET signal, whereas plasma pTau217 did not. The low discriminative ability of plasma pTau217 to identify early tau-PET stages aligns with previous findings.^31,32,70^ This observation is also consistent with the lower accuracy of pTau217 to detect AD pathology in CU compared to CI individuals^27,28^, which is likely due to lower pathology burden in CU than in CI individuals and argues against its stand-alone application in CU individuals.^71^ In contrast, VeraBIND Tau demonstrated similar diagnostic accuracies in both CI and CU individuals. While validation in independent cohorts is needed, these results suggest that the VeraBIND Tau assay is a highly promising, scalable tool to identify individuals with a high risk of clinical progression to AD, starting at early tau deposition stages (i.e., the entorhinal stage). This study also highlighted that the VeraBIND Tau assay may have affinity for detecting non-AD tauopathy, as results were positive in 77.8% of A-T+ individuals (n=7/9), including two A-T+ CU individuals and five A-T+ CI patients who were suspected to suffer from primary age-related tauopathy (PART, n=4) or fronto-temporal lobar degeneration (FTLD, n=1), as well as in a single A-T-case clinically suspected to suffer from cortico-basal degeneration, a 4R tauopathy. Including a larger number of cases with suspected PART may provide further insight into the performance of the VeraBIND Tau assay for identifying these individuals. Moreover, the diagnostic performance of the VeraBIND Tau assay in 4R tauopathies would need further investigation using radiotracers with higher affinity for 4R aggregates than [^18^F]MK6240.^59^

As pTau217 best predicted amyloid status, we tested whether a two-test approach combining low threshold plasma pTau217 (≥0.142pg/mL)^22^, as a Core 1 biomarker reflecting amyloid pathology^1^, and VeraBIND Tau assay, as a Core 2 biomarker reflecting tau pathology^1^, may help stratify individuals along the A/T continuum. In CU individuals, the A/T-like plasma stratification corresponded to the A/T CSF/PET classification in 83.5% of cases. Mismatches (16.5%) were mostly observed in individuals with discordant A/T CSF/PET classification (i.e., A-T+ and A+T-). Overall, this two-test approach identified all A+T+ individuals, while none of the CU individuals from the other A/T CSF/PET groups (i.e., A-T-, A+T-, A-T+) had positive results in this two-test approach. While validation is needed, these results show strong promise for screening CU individuals to support or rule out a presymptomatic AD diagnosis.^72^ Further studies in larger CU cohorts are warranted to assess the stand-alone and/or complementary value of the VeraBIND Tau assay to optimize first-line screening and reduce reliance on costly confirmatory examinations. In CI patients, concordant positive results on both assays were found for 91.5% of A+T+ individuals, while concordant negative plasma results were found in 100% of A-T-CI patients. These findings suggest that concordant positive results on both plasma tests are supportive of an AD etiology as the primary driver of cognitive impairment, while concordant negative results exclude this possibility. Discordant results, especially when only pTau217 is positive (>0.142pg/mL), should orient to confirmatory examinations. Future work will be necessary to investigate how to optimally use these plasma biomarkers and reduce reliance on lumbar puncture and/or PET imaging in larger and more diverse clinical populations.

Further longitudinal work is warranted prior to considering the use of VeraBIND Tau for monitoring the effects of anti-tau therapies. Both the cross-sectional VeraBIND Tau Score and its annual rate of change were associated with cross-sectional entorhinal tau-PET burden and Braak-like tau-PET stages, as well as with plasma pTau217 and pTau181 concentrations, cross-sectional and longitudinal episodic memory and MMSE scores. These findings suggest that VeraBIND Tau metrics are associated with markers of advancing AD pathology and cognitive decline. Nevertheless, plasma pTau217 appeared to exhibit a greater dynamic range across the Braak-like tau-PET spectrum than VeraBIND Tau (Fig. 3). Moreover, five participants (5.7%) converted to VeraBIND Tau positivity, whereas 83 of 88 (94.3%) maintained stable positive or negative results throughout follow-up, indicating high longitudinal classification stability. The absence of longitudinal tau PET data precluded determining whether conversion on VeraBIND Tau parallels conversion to tau-PET positivity. Ongoing follow-up will help characterize the biological and cognitive trajectories of the A-T-CU individuals who converted on VeraBIND Tau during follow-up. Together, these longitudinal findings suggest that VeraBIND Tau may provide a means of monitoring whether plasma pTau retains its ability to bind recombinant nTau over time, although its ability to dynamically track longitudinal changes in tau-PET signals needs further investigation.

Strengths of this study include head-to-head comparisons of VeraBIND Tau assay to several plasma biomarkers, well-characterized participants at the clinical and biological levels, and the inclusion of longitudinal plasma and cognitive data. The VeraBIND Tau assay has the advantage of relying on isolation and enrichment techniques that are compatible with the immunoassay format, which is easier to integrate in clinical workflow compared to mass spectrometry.

Limitations include a modest sample size, the absence of a validation cohort, and a relatively homogenous participant sample at the educational and racial levels, underscoring the need for replication in larger and more diverse cohorts. In addition, the CU individuals were enriched in APOE ɛ4 carriers, leading to an overrepresentation of individuals at elevated risk for developing AD^73,74^ relative to the general population. This enrichment may have inflated the assay diagnostic performance in this high-risk sample. Nevertheless, the diagnostic performance of VeraBIND Tau was directly compared to other plasma biomarkers within the same individuals. Further studies are warranted to evaluate its performance in larger, community-based cohorts. Moreover, there was an imbalance in the distribution of cases across Braak-like stage groups. Specifically, the number of individuals with Braak-like stages 1-3 (n=17) was low compared to those with stages 0 (n=76) or 4-6 (n=52). This distribution is consistent with the inclusion of symptomatic patients who are generally enriched in individuals with higher Braak-like stages and the low prevalence of early tau deposition, as assessed using tau-PET imaging, in CU samples.^6^ Nevertheless, this limited number of individuals with Braak-like stages 1-3 highlights the need to further investigate the value of VeraBIND Tau assay to detect pTau binding activity at the early stages of tau pathology in larger cohorts.

Despite these limitations, our study suggests that the VeraBIND Tau assay reliably captures early tau pathology regardless of amyloid status. Given its high accuracy for predicting tau-PET positivity, this assay could serve as a Core 2 plasma biomarker of underlying tau proteinopathy, as defined in the NIA-AA AD diagnostic framework ^1^. While further validation is needed, the VeraBIND Tau assay holds strong promise as a scalable and cost-effective alternative to tau-PET imaging for detecting tau pathology in CU individuals and for diagnosing symptomatic patients with suspected 3R/4R tau pathology (AD and PART). Moreover, our findings support that a two-test approach combining VeraBIND Tau with a low threshold pTau217 may constitute a valuable fluid-first workflow in a clinical setting and a powerful blood-based panel to accurately rule-in and rule-out AD pathology in CU individuals. Finally, our results highlight a potential utility of VeraBIND Tau for monitoring pTau binding activity in clinical trials.

## Data Availability

Dr. Hanseeuw has full access to all the data in the study and take responsibility for the integrity of the data and the accuracy of the data analysis. Request for data for replication studies or meta-analyses can be sent to the following email address.

## Acknowledgements

We thank the Stop Alzheimer’s Foundation for its support. We also thank Daniela Savina, Fiona Galande, and Florence Vanhoof for their help in the practical organization of the examinations, and Marine Van Calsteren, lab technician, for her contribution to the management of blood samples and SIMOA quantifications.

