## Supplementary Material for "Tau pathological binding activity in plasma before the onset of symptomatic Alzheimer’s disease"

### Materials and methods

#### APOE genotyping

All participants were analyzed for APOE polymorphisms rs429358 (which is a [C/T] substitution on chromosome 19q13.32 of the sequence GCTGGGCG CGGACATGGAGGACGTG[C/T]GCGGCCGCCTGGTGCAGTACCG CGG) and rs7412 (which is a [C/T] substitution of the sequence CCGCGATGCCGATGACCTGCAGAAG[C/T]GCCTGGCAGTGTACCAGGCCG GGGC). Based on the 2 single-nucleotide polymorphisms, 1 of the 3 alleles was assigned ε2, ε3, or ε4. Participants were classified into 2 groups: "ε4 carrier group" (ε2ε4, ε3ε4, ε4ε4) and "ε4 non-carrier group" (ε2ε2, ε2ε3 and ε3ε3).[73]

#### VeraBIND™ assay

Measurement of pathologically active hyperphosphorylated tau (PA pTau) in EDTA plasma was performed at Veravas, Inc. (805 Las Cimas Pkwy, Suite 150, Austin, TX 78746 USA, USA) and Access Genetics & OralDNA Labs (7400 Flying Cloud Drive; Eden Prairie, MN, 55344, USA) using the VeraBIND™ Tau plasma assay (Fig.1; Soldo, J., Iqbal, K., Bergmann, S., & Ansari, K. Detection of Disease State Macromolecules Binding to Normal Macromolecules as a Biomarker for Disease Identification. PCT/US24/50852. October 10, 2024). This assay was developed using the Veravas VeraBIND™ (Biomarker Isolation and eNrichment for Detection) sample transformation and biomarker purification technology (Soldo, J., Bergmann, S., & Wiley, C. Methods for Depletion and Enrichment. PCT/US2019/043711. July 27, 2019; Soldo, J., Bergmann, S., & Wiley C. Method for Detecting Biomarkers. PCT/US2019/043715. July 27, 2019; Soldo, J. Sample Depletion and Enrichment to Improve the Quality of Diagnostic Test Results. US Patent 10,948,484. March 16, 2021; Soldo, J., & Bergmann, S. System and Methods for Multiplex Detection of Biomarkers. PCT/US24/12072. January 18, 2024), a pool of proprietary monoclonal antibody-coated capture beads to specifically capture and purify pTau from plasma, supplied by ADx Neurosciences NV, A Fujirebio Company (Technologiepark 6, 9052 Gent, Belgium), recombinant full-length human Tau-441 with a V5 tag at the C-terminus expressed in HEK 293 cells supplied by Sigma-Aldrich, Inc. (PO Box 14508, St. Louis, MO 63178, USA), an anti-V5 tag monoclonal antibody conjugated to alkaline phosphatase (AP Conjugate) supplied by ThermoFisher (5781 Van Allen Way, Carlsbad, CA, 92008, USA), Lumi-Phos 530 AP Substrate supplied by Lumigen, Inc. (Substrate), a Beckman Coulter Company (22900 Eight Mile W, Southfield, MI, 48033, USA), the GloMax® Discover Microplate Reader supplied by Promega Corporation (2800 Woods Hollow Road, Madison, WI, 53711, USA), and the MINI 96 Pipette, 50 - 1250 µL (Part No. 4804) with Two Position Stage supplied by INTEGRA Biosciences Corp. (22 Friars Drive, Hudson, NH, 0305, USA). A TRIS-based sample diluent was prepared with a phosphatase inhibitor cocktail and nonionic surfactant to pre-analytically dilute and condition the freshly thawed 2-8°C EDTA plasma samples by irreversibly inactivating any endogenous phosphatases. TRU Block™ Ultra supplied by Meridian Bioscience (5171 Wilfong Road, Memphis, Tennessee, 38134, United States) was biotinylated and coupled to 1.6 µm VeraBIND streptavidin superparamagnetic beads and pooled with PEG-coated superparamagnetic beads to make custom VeraBIND clean beads to pre-analytically clean the diluted and conditioned EDTA plasma by selectively targeting and binding human anti-mouse antibodies (HAMA), rheumatoid factor, anti-PEG autoantibodies, anti-streptavidin autoantibodies, and non-specific binding interferences [67]. Different anti-hyperphosphorylated tau mouse monoclonal antibodies were biotinylated and coupled to the 1.6 µm VeraBIND streptavidin superparamagnetic beads to make a pool of custom VeraBIND capture beads to bind and purify any pTau in the diluted, conditioned and cleaned EDTA plasma. A TRIS-based Wash Buffer with nonionic surfactant was prepared to wash the capture beads and remove the EDTA plasma sample matrix, a TRIS-based Normal Tau Binding Buffer with nonionic surfactant was prepared for VeraBIND purified pTau to bind normal tau [33], and a Normal Tau Wash Buffer with nonionic surfactant was prepared to wash away excess recombinant normal tau and to wash away excess anti-V5 AP Conjugate without disrupting the [pTau]-[recombinant normal tau] PA pTau complex or aggregate.

Prior to starting the VeraBIND Tau assay protocol, the clean beads, capture beads, and freshly diluted recombinant V5-tagged normal tau (nTau) in normal tau bind buffer are equilibrated at room temperature on a laboratory mixer. The AD wash buffer and the Normal Tau binding buffer are equilibrated at Room temperature on the bench. The frozen EDTA plasma samples are thawed at 2-8°C in a cooler on a laboratory mixer for 60 minutes. 110 µL of freshly thawed 2-8°C EDTA plasma is added to 550 µL of 2-8°C assay-specific sample diluent in a 2 mL polypropylene (PP) micro tube (Sarstedt, Part No. 72.694.306) for 30 min at 2-8°C with gentle mixing to irreversibly inactivate any endogenous plasma phosphatases. 600 µL of the diluted and conditioned 2-8°C plasma sample is aspirated and dispensed into a 96 well non-sterile, PP 2mL deep well plate with round wells and round bottom (Stellar Scientific, Part No. DWP-WB-3866) with 100 µL custom VeraBIND clean beads to pre-analytically clean the sample for 30 min at 37°C and 1,000 rpm on a plate heater/shaker (Qinstruments BioShake iQ with plate-specific thermal adapter, Part No. 1808-0506). After the clean beads are magnetically separated on a plate magnet (Alpaqua Magnum FLX® with Solid-Core™ Technology) for 15 min, 700 µL bead-free sample supernatant is aspirated and dispensed into a new 96 well non-sterile, PP 2mL deep well plate with round wells and round bottom with 100 µL custom VeraBIND capture beads. The capture beads and sample are mixed for 30 min at 37°C and 1,000 rpm on a plate heater/shaker for the selective capture and purification of any hyperphosphorylated tau (pTau) by the pool of monoclonal antibody-coated capture beads. After the incubation, the capture beads are magnetically separated on a plate magnet (Alpaqua Magnum FLX® with Solid-Core™ Technology) for 9 min, and the supernatant aspirated and discarded. The capture beads are magnetically washed 4x with 500 µL AD wash buffer to remove the plasma matrix, and then the capture beads are buffer exchanged, and pipette mixed into 120 µL normal tau binding buffer to facilitate ionic and hydrophobic binding of PA pTau to recombinant normal tau (nTau). Next, 100 µL of recombinant V5-tagged full-length normal tau 1-441 (Recombinant Normal Tau-441) is added, pipette mixed and incubated static (no mixing) overnight in a 2-8°C cooler for the binding of the Recombinant Normal Tau-441 by any PA pTau, akin to PA pTau mediated normal tau aggregation observed in the brain of AD patients. The next day, normal tau wash buffer, detection buffer, and substrate are equilibrated at room temperature, the anti-V5 AP Conjugate is freshly diluted, and freshly thawed TruBlock Ultra is added to the detection buffer. After the capture beads are warmed to 37°C for 30 min and magnetically washed 4x with 500 µL normal tau wash buffer to remove any excess or non-bound Recombinant Normal Tau-441, 100 µL of detection buffer with Tru Block Ultra is added, pipette mixed and incubated for 5 minutes. 100 µL of the freshly diluted AP Conjugate is added, pipette mixed and incubated static (no mixing) at 37°C for 30 min to detect any Recombinant Normal Tau-441 bound by PA pTau on the capture beads. After magnetically washing the capture beads 4x with 500 µL normal tau wash buffer to remove any excess or non-bound AP Conjugate, the capture beads are pipette mixed with 200 µL normal tau wash buffer, aspirated and dispensed into a Corning® 96-well white round bottom polystyrene NBS Microplate (Corning, Part No. 3605), magnetically separated on the plate magnet, and the supernatant aspirated and discarded. 100 µL substrate is added, mixed for 60 seconds at 1000 rpm on the heater/shaker, and incubated for 60 min at 30°C static (no mixing) to generate a luminescence signal that is read by a GloMax® Discover Microplate Reader. The relative luminescence units (RLU) generated by the substrate is directly proportional to the amount of Recombinant Normal Tau-441 bound by PA pTau and serves as a representation of the PA pTau captured and purified from the plasma sample.

The semi-quantitative VeraBIND Tau assay results are reported as test result Score which is calculated using an EDTA plasma-based Standard (e.g., EDTA plasma collected from patients negative for tau pathology by [$^{18}$F]MK6240 tau-PET imaging, or EDTA plasma collected from apparently healthy adults age 18-32). The Standard is run in duplicate in each assay, and each lot of Standard has a lot-specific correction factor which is used to set the assay cutoff RLU for each run. The test result Score for each sample is calculated by dividing the patient sample test result (RLU) by the assay cutoff RLU, or the test result Score = [Unknown signal response (RLU)] / [(Standard test result signal (RLU))*(Correction Factor)]. A test result Score <1.0 is a negative test result, meaning that PA pTau has not been detected in the plasma sample, while a test result Score ≥1.0 is a positive test result, indicating that PA pTau has been detected in the plasma sample. The VeraBIND Tau assay Negative Control is a pool of EDTA plasma collected from patients negative for tau pathology by [$^{18}$F]MK6240 tau-PET imaging or EDTA plasma collected from apparently healthy adults age 18-32 with a test result Score <0.93. The VeraBIND Tau assay Positive Control is a pool of EDTA plasma collected from patients positive for tau pathology by [$^{18}$F]MK6240 tau-PET imaging with a test result Score >1.20.

#### Amyloid-PET acquisition and reconstruction procedures

For the [$^{18}$F]Flutemetamol PET-CT, a 30-minutes list-mode acquisition was performed on a Philips Gemini PET (Philips Healthcare, Amsterdam, Netherlands) 90 minutes after intravenous injection (target activity 185±5 MBq). The images were reconstructed as a dynamic scan of 6×5 minutes frames with 2 mm isometric voxels including attenuation, scatter and decay corrections, and time-of-flight information using the manufacturer's standard reconstruction algorithm.

For the [$^{11}$C]PiB PET-CT, a 20 min list-mode acquisition was performed on a Philips Vereos digital PET (Philips Healthcare, Amsterdam, Netherlands) forty minutes after intravenous injection (target activity 500MBq). Images were reconstructed in 4 x 5 minutes frames with 2 mm isometric voxels using the manufacturer's reconstruction algorithm which includes attenuation, scatter, and decay corrections, and time-of-flight information using the manufacturer's standard reconstruction algorithm.

#### Visual Braak-like staging

Braak-like ROIs were defined based on previous work led by Schöll et al. on in vivo Braak staging using brain imaging.[47] Braak-like stage 0 corresponded to the absence of elevated radiotracer uptake. Braak-like stages 1–2 were defined by tracer retention restricted to the medial temporal lobe (i.e., entorhinal region, hippocampus, parahippocampal gyrus). Braak-like stages 3–4 indicated involvement of the temporal neocortex and/or posterior cingulate cortex. Braak-like stage 5 was assigned when tracer uptake was detected in the frontoparietal neocortex and/or the occipital lobe, whereas Braak-like stage 6 corresponded to significant radiotracer uptake in precentral, postcentral, paracentral gyri, cuneus and pericalcarine regions.[45]

### Supplementary Table 1

**Supplementary Table 1. Linear mixed-effects model results for the association between VeraBIND Tau scores and longitudinal decline in MMSE and episodic memory composite scores, adjusted for age, sex, and education, with and without additional adjustment for plasma pTau217 concentration.**

| | β | SE | 95% Conf. Interval | t | p |
|---|---|---|---|---|---|
| **MMSE** | | | | | |
| (Intercept) | 19.41 | 4.86 | 9.84 – 28.97 | 3.99 | **<0.001** |
| Time (years) | 3.04 | 0.46 | 2.13 – 3.94 | 6.60 | **<0.001** |
| VeraBIND Tau score (RLU ratio) | -1.05 | 1.78 | -4.56 – 2.45 | -0.59 | 0.555 |
| Age (years) | 0.04 | 0.05 | -0.07 – 0.15 | 0.75 | 0.451 |
| Sex (Male) | -2.03 | 0.88 | -3.76 – -0.30 | -2.31 | **0.021** |
| Education (years) | 0.42 | 0.13 | 0.17 – 0.67 | 3.25 | **0.001** |
| Time x VeraBIND Tau score | -3.59 | 0.47 | -4.52 – -2.66 | -7.63 | **<0.001** |
| **MMSE (adjust. for pTau217)** | | | | | |
| (Intercept) | 21.74 | 3.89 | 14.07 – 29.40 | 5.59 | **<0.001** |
| Time (years) | 3.03 | 0.46 | 2.12 – 3.93 | 6.60 | **<0.001** |
| VeraBIND Tau score (RLU ratio) | 3.06 | 1.57 | -0.04 – 6.16 | 1.94 | 0.053 |
| Plasma pTau217 concentration (pg/mL) | -6.63 | 0.89 | -8.39 – -4.87 | -7.42 | **<0.001** |
| Age (years) | 0.01 | 0.04 | -0.08 – 0.09 | 0.13 | 0.899 |
| Sex (Male) | -1.27 | 0.70 | -2.65 – 0.11 | -1.81 | 0.072 |
| Education (years) | 0.30 | 0.10 | 0.09 – 0.50 | 2.86 | **0.005** |
| Time x VeraBIND Tau score | -3.58 | 0.47 | -4.50 – -2.66 | -7.63 | **<0.001** |
| **Episodic memory z-score** | | | | | |
| (Intercept) | 3.56 | 2.30 | -0.98 – 8.10 | 1.55 | 0.124 |
| Time (years) | 0.60 | 0.24 | 0.13 – 1.07 | 2.54 | **0.012** |
| VeraBIND Tau score (RLU ratio) | -5.74 | 1.23 | -8.17 – -3.31 | -4.65 | **<0.001** |
| Age (years) | -0.02 | 0.02 | -0.07 – 0.03 | -0.78 | 0.436 |
| Sex (Male) | -0.40 | 0.39 | -1.16 – 0.36 | -1.04 | 0.299 |
| Education (years) | 0.16 | 0.06 | 0.04 – 0.27 | 2.72 | **0.007** |
| Time x VeraBIND Tau score | -0.73 | 0.25 | -1.23 – -0.23 | -2.89 | **0.004** |
| **Episodic memory z-score (adjust. for pTau217)** | | | | | |
| (Intercept) | 3.64 | 2.03 | -0.36 – 7.63 | 1.79 | 0.074 |
| Time (years) | 0.61 | 0.24 | 0.15 – 1.08 | 2.60 | **0.010** |
| VeraBIND Tau score (RLU ratio) | -2.90 | 1.23 | -5.33 – -0.48 | -2.36 | **0.019** |
| Plasma pTau217 concentration (pg/mL) | -2.41 | 0.48 | -3.36 – -1.47 | -5.06 | **<0.001** |
| Age (years) | -0.04 | 0.02 | -0.08 – 0.01 | -1.70 | 0.090 |
| Sex (Male) | -0.15 | 0.34 | -0.83 – 0.52 | -0.44 | 0.658 |
| Education (years) | 0.10 | 0.05 | 0.00 – 0.21 | 1.97 | 0.051 |
| Time x VeraBIND Tau score | -0.75 | 0.25 | -1.25 – -0.25 | -2.97 | **0.003** |

Legend. Conf. Interval = confidence interval; adjust. = adjustment; RLU = relative luminescence signal; pg = picogram; mL = milliliter.

### Supplementary Figures

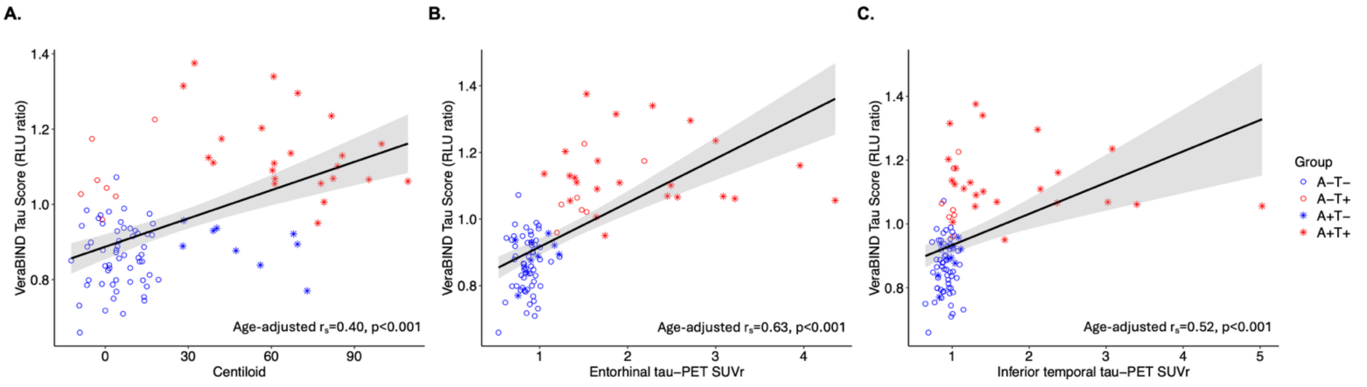


**Supplementary Fig.1.** Associations between VeraBIND Tau semi-quantitative measure (relative luminescence unit [RLU] ratio) and: (**A)** Centiloid value, (**B)** Entorhinal tau burden, as measured using [$^{18}$F]MK6240 tau-PET Standard Uptake Value ratio (SUVr), **(C)** Inferior temporal tau burden (SUVr), in the subsample of 96 participants with both available amyloid-PET and tau-PET data.

A- = amyloid negative; A+ = amyloid positive; CU = cognitively unimpaired; CI = cognitively impaired.

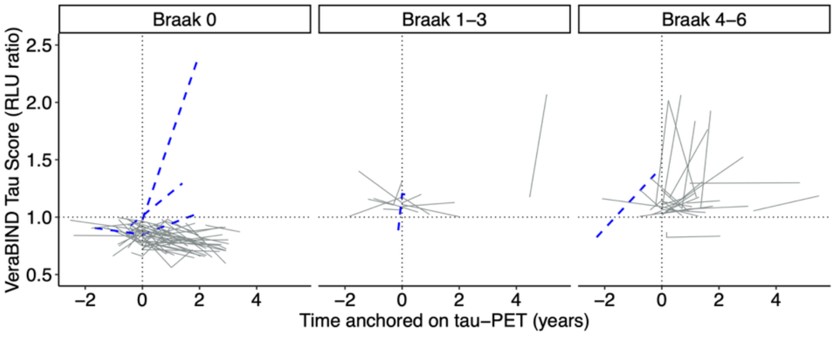

**Supplementary Fig. 2.** Longitudinal VeraBIND Tau scores (relative luminescence unit [RLU] ratio) in Braak-like tau-PET stage groups. The blue dashed lines represent individuals who converted from a negative test result (RLU ratio score < 1.0) to a positive test result (RLU ratio score ≥ 1.0).
